# Local and circulating cytotoxic CD4⁺ T cells are early markers of disease activity in pediatric Crohn’s disease

**DOI:** 10.64898/2026.06.16.26354238

**Authors:** Veronika Niederlova, Kyle Kimler, Hengqi Betty Zheng, Marc Elosua Bayes, Rose Hedderman, Paula Keskula, Iva Pacakova, Jan Vecek, Joshua de Sousa Casal, Andrew Kwong, Leonard Nettey, Zoë Steier, Katarína Kováčová, Brandi Bratrude, Jillian Zavistaski, Wei Keat Lim, Andrea T. Hooper, Scott MacDonnell, Nathalie Fiaschi, Zaruhi Hovhannisyan, Ghassan Wahbeh, David Suskind, Lusine Ambartsumyan, Dale Lee, Jan Dobeš, Scott B. Snapper, Ondrej Stepanek, Alex K. Shalek, Leslie S. Kean, Jose Ordovas-Montanes

## Abstract

Inflammatory bowel disease (IBD) burden is rising globally, yet only subsets of patients benefit from available therapies, underscoring the need for more precise molecular and cellular stratification. In the PREDICT study, we enrolled treatment-naïve pediatric patients with IBD, alongside disorders of gut–brain interaction (DGBI) controls and healthy donors, and profiled their intestinal and blood-derived T cells using single-cell RNA sequencing (scRNA-seq). Across 107 participants, we identify a unique population of cytotoxic CD4⁺ T cells (CD4 CTL) enriched in the inflamed gut of patients with Crohn’s disease (CD) and ulcerative colitis. CD4 CTLs are clonally expanded and express cytotoxic effector molecules and *IFNG*, consistent with antigen-driven activation. Cell–cell interaction analyses implicate macrophage-derived IL-27 as the top candidate for CD4 CTL differentiation, and IL-27 blockade in a mouse model limits CD4 CTL formation. Notably, elevated CD4 CTL frequencies in gut and peripheral blood at diagnosis are associated with subsequent poor outcome of anti-TNF therapy in pediatric CD. Findings in our identification cohort are validated in an independent cohort and through reanalysis of published datasets. Importantly, we designed a simple flow cytometry panel to isolate blood CD4^+^ CXCR6^+^ CD27^-^ T cells, which displayed a CD4 CTL transcriptional phenotype. Together, our results link CD4 CTLs to anti–TNF nonresponse and support their potential as an early, blood-accessible biomarker for treatment stratification in pediatric CD.

## Introduction

Inflammatory bowel disease (IBD), which includes Crohn’s disease (CD) and ulcerative colitis (UC), is a group of chronic immune-mediated disorders of the gastrointestinal (GI) tract with increasing global prevalence^1^. Although the range of available biologic therapies to complement TNF inhibitors is improving disease management, up to 50% of patients continue to exhibit primary nonresponse^2,3^. Currently, there are no reliable biomarkers to predict treatment outcomes at diagnosis, making personalized treatment strategies with approved anti-TNFs, anti-IL-12/23 pathway inhibitors, anti-α4β7 inhibitors, and JAK inhibitors difficult to implement. An ideal cellular biomarker would be readily detectable in blood, connected to immunopathology in the gut, and provide an actionable set of potential targets.

CD4⁺ T cells are central players in IBD pathogenesis. Genome-wide association studies have repeatedly implicated genes involved in antigen presentation and T cell regulation, including HLA class II alleles and T cell polarizing cytokine receptors such as *IL23R*, highlighting aberrant CD4⁺ T cell responses as key genetic underpinnings of disease^4–6^. Functionally, CD4⁺ T cells orchestrate mucosal inflammation through interactions with antigen-presenting cells, cytokine secretion, and recruitment of other effector cells, such as neutrophils and inflammatory monocytes^7–10^. These functions depend on proper differentiation of T helper subsets, which is often imbalanced in IBD, contributing to both excessive inflammation and loss of immune tolerance^11,12^. Therapeutic targeting of TNF, IL-23 pathway, and JAK/STAT signaling further reinforces the concept that T cell balance is critical to maintain gut homeostasis. Unlike classical autoimmune diseases, the antigens driving T cell activation in IBD span the range from commensal bacteria or fungi to viral and self-antigens, leading to a broader autoinflammatory classification^13–16^. Despite their central role, the full heterogeneity, origins, and disease-specific functions of CD4⁺ T cells in the human gut remain incompletely understood.

Single-cell RNA sequencing (scRNA-seq) has recently enhanced our understanding of intestinal immune dysregulation in IBD^4,17–21^. Prior studies have mapped cellular networks and identified pathogenic cell states associated with poor treatment outcomes, such as IL13RA2⁺IL11⁺ inflammatory fibroblasts in UC^22^, inflammatory macrophages, activated dendritic cells, plasma cells, and stromal cells in CD^5^, as well as protective subsets like *TREM2*-expressing *C1Q*^hi^ *IL1B*^lo^ macrophages associated with CD remission^6^. In the same study, analysis of samples from adult patients with CD and UC before and after anti-TNF blockade highlighted IFN-response signatures in T cell aggregates around epithelial damage in CD and UC, with a further Th1 polarization in CD^6^. Our own work focusing on distinct colonic pathology in CD and UC from pediatric patients reinforces the Th1- and Th17-associated T cell activation programs that correlate with anti-TNF response^23^. However, although the Th17/IL-23 axis has yielded tractable therapeutic targets, the specific effector states and pathways underlying Th1-associated pathology in CD remain less clearly defined. A more refined understanding of pathogenic CD4⁺ T cell states linked to treatment outcome may therefore help identify actionable biomarkers and therapeutic targets^24,25^.

To address this need, we conducted the PREDICT study, integrating scRNA-seq of intestinal biopsies and peripheral blood mononuclear cells (PBMC) from treatment-naïve pediatric IBD patients, patients with disorders of gut-brain interaction (DGBI), and healthy donors. Here, we identify a cytotoxic CD4⁺ T cell subset (CD4 CTL) enriched in both intestinal tissue and blood of CD patients, particularly those with poor response to anti-TNF therapy. These CD4 CTLs appear to originate from antigen-driven clonal expansion and cytokine-mediated programming, implicating IL-27, IFN-γ and type I interferons as candidate polarizing signals. Importantly, we identify shared TCR clonotypes between blood and inflamed tissue. In addition to their potential as an early blood-based biomarker for anti-TNF nonresponse, these findings nominate CD4 CTLs and their associated pathways as candidate therapeutic targets in refractory CD.

## Results

### Patient cohort and study design

The PREDICT study enrolled pediatric participants presenting with GI symptoms suggestive of IBD prior to diagnostic workup and treatment initiation (**Fig. 1A**). The complete profiled PREDICT cohort included 107 participants. Our complete analysis cohort comprised 43 treatment-naïve patients between 6 and 19 years old (median 12) diagnosed with CD, 19 with UC (7–19 years old, median 13), and 28 individuals without evidence of intestinal inflammation who were ultimately diagnosed with DGBI (6–21 years old, median 15). DGBI patients, who exhibit GI symptoms in the absence of overt macroscopic mucosal inflammation or autoimmunity, served as the primary control group for characterization of the T-cell compartment. To further support our analysis, we also included 9 healthy young adult donors, acknowledging their older age (18–24 years old, median 23) due to the ethical limitations of collecting intestinal samples from healthy children. Eight participants classified as other diagnoses were included in atlas generation and quality-control analyses but were excluded from disease-group statistical comparisons. Complete clinical metadata for all participants is provided in **Supplementary Table 1** and in **Fig. S1A-C**.

**Figure 1.**
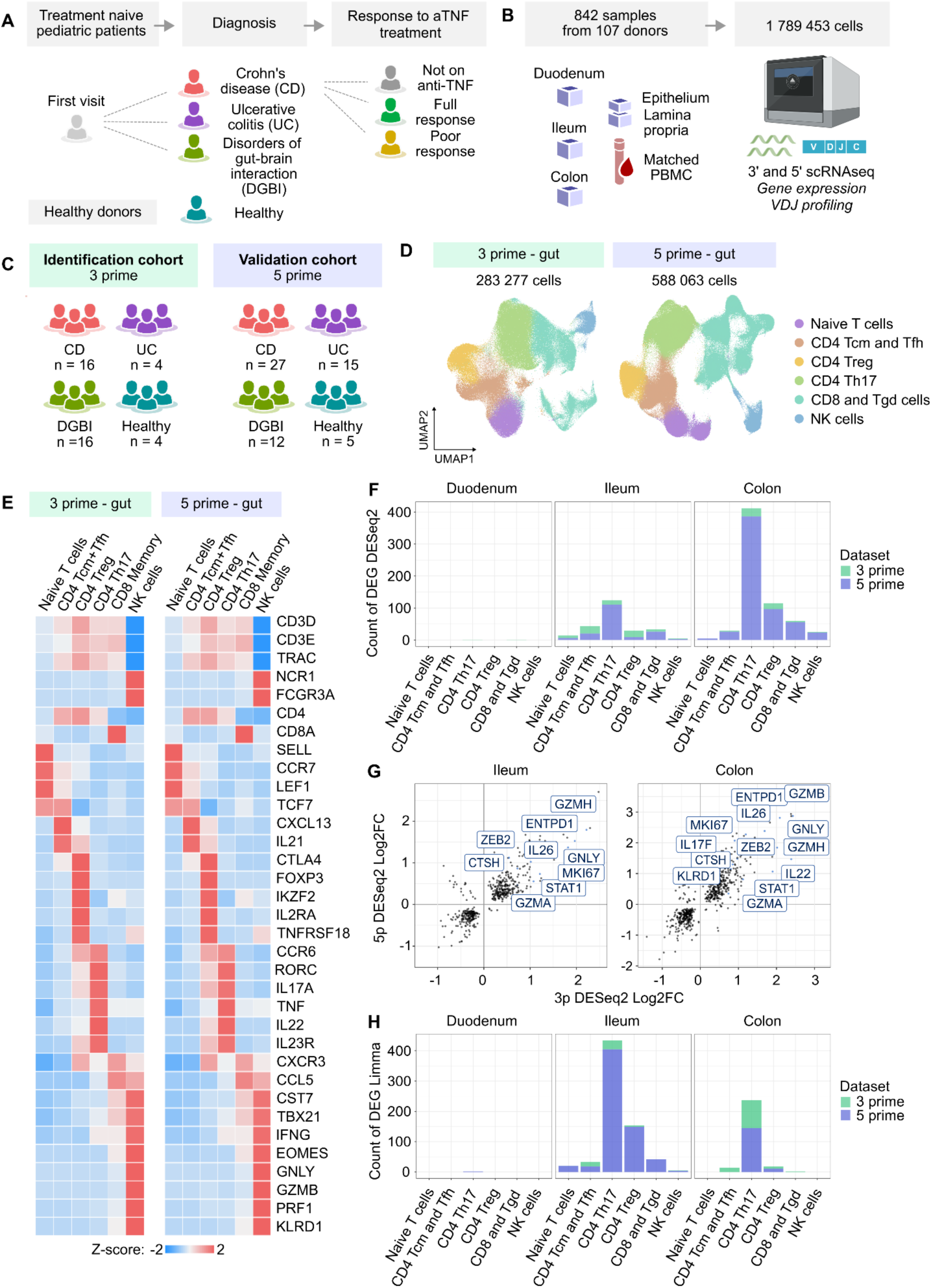
Analysis of T cells and NK cells from the PREDICT cohort by scRNA-seq. (A–C) Overview of the cohort (A), analyzed samples (B), and allocation to the identification and validation cohorts (C). **(A)** The PREDICT study enrolled treatment-naïve pediatric patients presenting with gastrointestinal symptoms. Based on diagnostic criteria, participants were classified as Crohn’s disease (CD), ulcerative colitis (UC), or disorders of gut-brain interaction (DGBI). CD participants were further stratified by response to anti-TNF therapy during two-year follow-up. Healthy controls were young adult volunteers and were not age-matched to the pediatric cohort. **(B)** T cells from 842 gut and blood samples obtained from 107 participants were profiled. Biopsies from duodenum, ileum, and colon, as well as PBMC, were collected at diagnosis and processed using 10x Genomics 3p or 5p chemistry. Epithelial and lamina propria fractions were separated enzymatically. **(C)** The identification cohort comprised samples from 40 donors processed with 10x 3p sequencing; the validation cohort included samples from 59 donors processed with 10x 5p sequencing. Donors represented in both datasets contributed to statistical analyses only in the identification cohort. **(D)** UMAP projections of T lymphocytes from the gut PREDICT 3p dataset (n = 283,277 cells from 40 donors) and the gut PREDICT 5p dataset (n = 588,063 cells from 99 donors, including the 40 donors from the 3p dataset). Cells are colored by manual annotation. **(E)** Heatmaps showing selected marker genes characterizing the clusters in (D) for the 3p dataset (left) and the 5p dataset (right). Colors indicate row-scaled z-scores of average gene expression per cluster. **(F–H)** Pseudobulk analyses were performed by aggregating counts per cluster, participant, and tissue, followed by differential expression analysis using DESeq2 to compare IBD (CD + UC) versus non-IBD (DGBI + healthy) samples. **(F)** Number of differentially expressed genes between IBD and non-IBD samples identified in the 3p and 5p datasets. **(G)** Volcano plots showing selected differentially expressed genes with the highest log2 fold-change differences between IBD and non-IBD samples in the CD4+ Th17 cluster from the 3p and 5p datasets. **(H)** Bar plot shows the counts of DEG in each T-cell and NK-cell population whose expression significantly correlated with anti-TNF treatment outcome in participants with CD. Differential expression was assessed using Limma with treatment response modeled as an ordinal variable (0 = not on anti-TNF, 1 = full response, 2 = partial response). DEG represent those with significant expression changes associated with progressively poorer treatment outcome in the 3p and 5p datasets.

Patients were followed for at least two years after initial endoscopy. Within the CD group, we stratified patients into three categories: those who did not require biologics (“not on anti-TNF”, NOA), those who achieved sustained remission on anti-TNF therapy (“full response”, FR), and those with partial response (PR).

At diagnosis, we collected biopsies from the duodenum, terminal ileum, and colon, alongside PBMCs of the same patients. Biopsies were separated into epithelial and lamina propria (LP) fractions, yielding a total of 842 samples from 107 donors (**Fig. 1B, Fig. S1A, Supplementary Table 1**). In 13 donors, we obtained repeated samples after the initiation of treatment.

### Single-cell profiling and data integration

To characterize T-cell populations in IBD, we initially analyzed a subset of 40 participants, termed the “**identification cohort**,” using the 10x Genomics 3’ scRNA-seq platform (“3p dataset”). To confirm the robustness of our findings, we analyzed additional samples from an independent set of 59 donors (the “**validation cohort**”) using the 10x Genomics 5’ platform (“5p dataset”), which also enabled TCR repertoire analysis (**Fig. 1C**). For a subset of donors (n = 36), we profiled the same sample using both 3p and 5p chemistries, allowing us to assess technical consistency. In such cases, donors were assigned exclusively to the identification cohort for statistical evaluation.

First, we focused on analysis of the gut samples. In total, 283,277 T and NK cells passed quality control (QC) in the 3p dataset, and 588,063 cells passed QC in the 5p dataset (**Fig. 1D**). We applied STACAS-based integration^26^ over variables 10x version, sample batch and donor ID to correct for batch effects while preserving true biological variation between tissue regions and disease status (**Fig. S1D**). The main source of variability persisting between samples post-integration was the layer of origin of lymphocytes – LP versus epithelial layer – reflecting anticipated biology (**Fig. S1D-E**).

Unsupervised clustering identified canonical T and NK cell subsets across both 3p and 5p datasets (**Fig. 1E**). Naïve T cells expressed high levels of *SELL*, *CCR7*, *LEF1*, and *TCF7*. Non-naïve CD4⁺ T cells included central memory (Tcm) distinguished by the downregulation of naïve markers, and follicular helper (Tfh) subsets expressing *CXCL13* and *IL21*. CD4^+^ Th17 cells were characterized by high expression of *RORC* (encoding transcription factor RORγt), *IL17A*, *CCR6* and *IL23R*. CD8⁺ memory T cells co-expressed *CXCR3*, *CCL5*, *CST7*, *TBX21*, and *EOMES*. NK cells were characterized by expression of *NCR1*, *KLRD1*, *FCGR3A* (CD16), *GNLY*, *GZMB*, and *PRF1*, and lacked T cell receptor expression. The relative frequencies of these cell types were highly correlated across the 3p and 5p methods, confirming methodological consistency (**Fig. S2A**).

### CD4⁺ Th17 cell remodeling associates with IBD and anti-TNF treatment outcome

To identify transcriptional differences between conditions, we performed pseudo-bulk differential expression analysis using DESeq2^27^ in the main populations of T cells and NK cells. For initial comparisons, we grouped CD and UC patients into a single IBD category and compared them to non-IBD controls (DGBI and healthy donors). Across cell types, the 5p dataset yielded a larger number of statistically significant differentially expressed genes (DEG) than the 3p dataset, consistent with increased statistical power driven by its larger cohort size (**Fig. 1F**, **Fig. S2B**). Despite these differences in DEG counts, differential expression patterns were highly concordant between the 3p and 5p datasets, particularly in LP samples from ileum and colon, supporting the robustness of the approach and indicating that the observed signatures were not driven by cohort- or chemistry-specific effects (**Fig. S2C**).

The largest number of DEGs was observed in the CD4⁺ Th17 subset, suggesting pronounced transcriptional remodeling of this subset in IBD (**Fig. 1F**). Among the most strongly upregulated genes in IBD CD4⁺ Th17 cells were those associated with proliferation (*MKI67*), immunoregulation (*ENTPD1*, encoding CD39), interferon signaling (*STAT1*), and cytotoxicity (*GZMH*, *GNLY*, *GZMB*) (**Fig. 1G**). The acquisition of cytotoxic effector gene expression within the Th17 lineage is atypical, and points to the emergence of a cytotoxic program in CD4⁺ T cells in IBD, consistent with emerging literature^18,20^. When comparing CD patients stratified by anti-TNF treatment outcome, the CD4⁺ Th17 subset again showed the highest number of DEGs meeting statistical significance (**Fig. 1H**). This suggests that transcriptional variation within this subset may contain clinically actionable biomarkers linked to therapeutic response.

Together, these analyses reveal that CD4⁺ Th17 cells undergo profound inflammatory and cytotoxic reprogramming in pediatric IBD. While prior evidence has been limited^28,29^, this observation supports the Th17 compartment as a central node in disease pathogenesis and treatment outcome.

### Identification of cytotoxic CD4⁺ T cells in IBD

Given the pronounced transcriptional differences in CD4⁺ Th17 cells between IBD and non-IBD samples, we performed a focused reanalysis of CD4⁺ T cell subsets in both the 5p (**Fig. 2A**) and 3p (**Fig. S3A**) datasets. Subclustering revealed 11 transcriptionally distinct CD4⁺ T cell subsets, which we annotated based on clonal expansion rate (**Fig. 2B**), gene expression profiles (**Fig. 2C**, **Fig. S3B**), and tissue localization (**Fig. 2D-E**, **Fig. S3C-D**).

**Figure 2.**
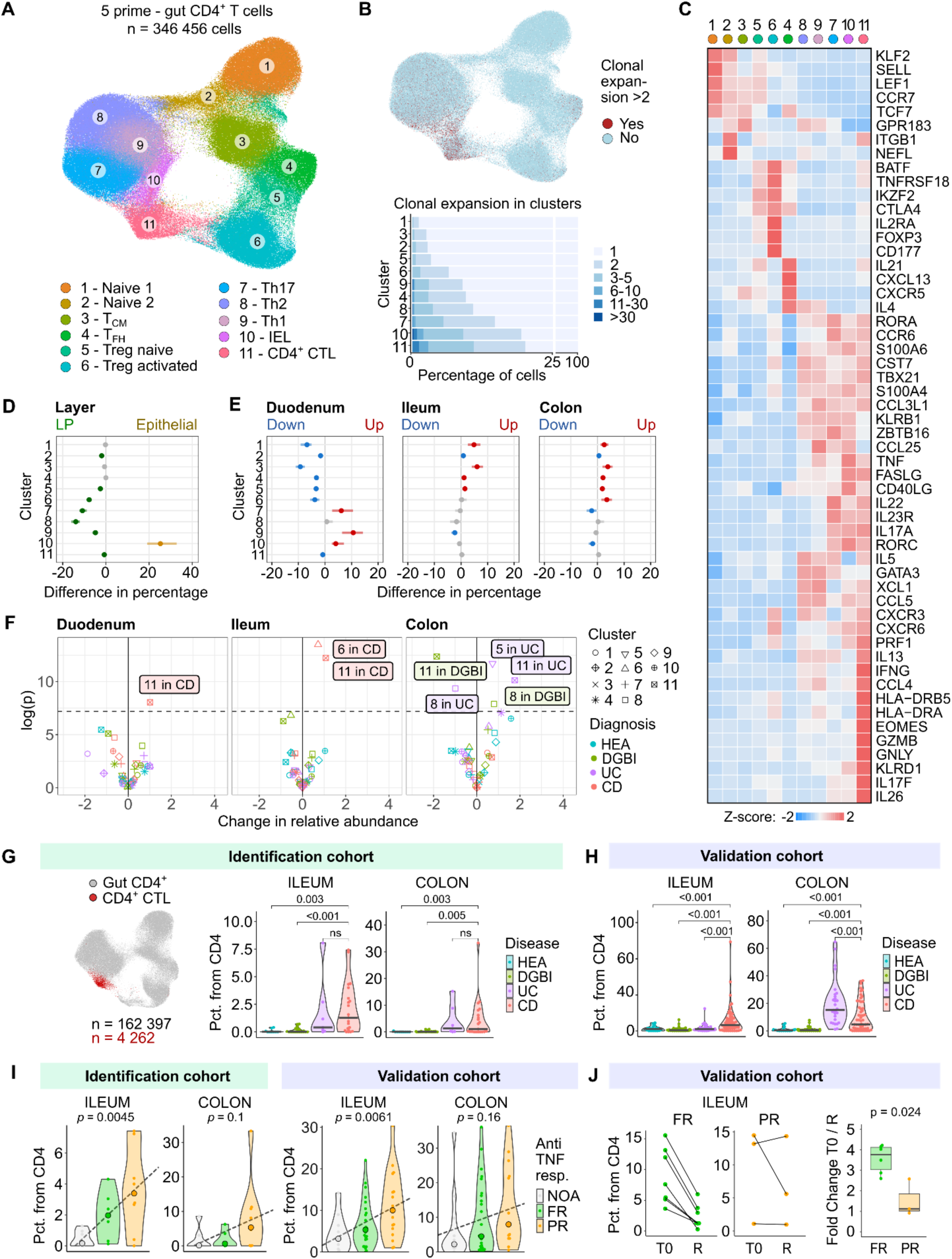
CD4⁺ T cells in the gut of IBD participants form a cytotoxic CD4 CTL population. **(A)** UMAP projection of CD4⁺ T lymphocytes from the gut PREDICT 5p dataset (n = 346,456 cells from 99 donors). Cells are colored according to manual cluster annotations. **(B)** Analysis of TCR repertoires within CD4⁺ T-cell populations. *Top*: UMAP projection as in (A), with cells colored by clonal expansion status. Expansion is calculated as the log₂ frequency of identical TCR CDR3α and CDR3β sequence pairs in the dataset. *Bottom*: Quantification of clonal expansion across the clusters shown in (A). **(C)** Heatmap of selected marker genes defining the clusters shown in (A). Colors represent row-scaled z-scores of average gene expression within each cluster. **(D–E)** Forest plots showing changes in cluster proportions across intestinal compartments. Comparisons are shown between lamina propria (LP) and epithelium (D), and across different gut locations (E). **(F)** Volcano plot showing relative changes in the proportions of subsets from Fig. 2A in patients with Crohn’s disease (CD), ulcerative colitis (UC), DGBI, and healthy donors from the PREDICT 5p gut dataset. Subset proportions were subjected to centered log-ratio normalization. P values were calculated using two-tailed Mann–Whitney U tests with Bonferroni correction for multiple comparisons. **(G)** Proportions of CD4 CTL cells in participants from the identification cohort profiled using 3p technology. *Left*: UMAP projection of CD4⁺ T cells from the 3p dataset. *Right*: Violin plots showing the percentage of CD4 CTL cells among total CD4⁺ T cells. P values were calculated using a two-tailed Mann–Whitney U test with Bonferroni correction for multiple comparisons. Horizontal bars indicate medians. **(H)** Proportions of CD4 CTL cells in participants from the validation cohort profiled using 5p technology. Violin plots show the percentage of CD4 CTL cells among total CD4⁺ T cells. P values were calculated as in (F). Horizontal bars indicate medians. **(I)** Proportions of CD4 CTL cells in participants with CD from the identification cohort (*left*) and validation cohort (*right*), stratified by response to anti-TNF therapy. NOA, not on anti-TNF treatment; FR, full response; PR, partial response. P values were calculated using two-tailed Pearson correlation tests. Medians are indicated by large dots. **(J)** Proportions of CD4 CTL cells in participants with CD from the validation cohort stratified by treatment response in different sampling timepoints. T0 – timepoint at diagnosis, R – repeated sampling in the same donor after 3 months. On right, quantification of the fold changes between percentages at T0 and R. P-value calculated using two-tailed Mann Whitney U test.

Several clusters represented naïve or early memory subsets: naïve CD4⁺ T cells (clusters 1 and 2), CD4⁺ Tcm (cluster 3) and naïve T regulatory (Treg) cells (cluster 5). These subsets showed a low clonal expansion (**Fig. 2B**) and expressed canonical naïve/memory markers such as *CCR7* and *LEF1*, with *KLF2* and *SELL* restricted to naïve cells. Activated Tregs (cluster 6) were identified by expression of *FOXP3*, *IL2RA* (CD25), *TNFRSF18* (GITR), and *IKZF2* (HELIOS). T follicular helper (Tfh) cells (cluster 4) expressed *CXCR5*, *CXCL13*, and *IL21*. Th2 cells (cluster 8) were characterized by *GATA3*, *IL4* and *IL5* and Th1 cells (cluster 9) expressed *TBX21*, *CXCR3* and *TNF*.

Three clusters (clusters 7, 10, and 11) shared hallmark Th17-associated genes (*IL17A*, *IL23R*, *CCR6*, *RORC*), but exhibited distinct phenotypes. Cluster 7 represented classical Th17 cells, cluster 10 corresponded to intraepithelial lymphocytes (IEL) and cluster 11 consisted of highly activated Th17-like cells expressing *PRF1*, *IFNG*, *GZMB*, *GNLY*, and *HLA-DRA*, suggesting acquisition of cytotoxic function (**Fig. 2C**). Gene set enrichment analysis (GSEA) confirmed a cytotoxic program in cluster 11 (**Fig. S3E**). Based on these features, we identified this subset as CD4⁺ cytotoxic lymphocytes (CD4 CTL).

We assessed cluster enrichment by layer (epithelial vs. LP) showing enrichment of cluster 10 IELs in epithelial layers and relative enrichment of classical cluster 7 Th17 and cluster 8 Th2 cells in LP layers (**Fig. 2D**). We then assessed cluster enrichment by intestinal region (e.g. duodenum vs. rest, etc.) and identified relative enrichment of cluster 9 Th 1 cells in duodenum relative to other regions (**Fig. 2E**).

We hypothesized that the cluster 11 CD4 CTL population accounted for the cytotoxic gene expression signature observed earlier in pseudo-bulked Th17 cells in IBD patients (**Fig. 1G**). To test this, we performed proportionality analysis using center log-ratio transformation of cell cluster frequencies across samples^30^. CD4 CTLs were significantly enriched in the inflamed gut of CD patients, particularly in the duodenum and ileum, and in the colons of UC patients (**Fig. 2F**), with consistent results across both identification and validation cohorts (**Fig. 2G-H**).

### CD4 CTLs are reproducibly detected across cohorts and linked to anti-TNF response

To determine whether CD4 CTLs were also present in previously published datasets, we reanalyzed two publicly available datasets: a pediatric CD cohort^21^ and an adult IBD cohort comprising both CD and UC patients pre- and post-biologic initiation^6^.

In both datasets, we extracted and reclustered CD4⁺ T cells (**Fig. S4A-B**). In each cohort, we identified a distinct cluster of CD4⁺ T cells expressing the same cytotoxic gene program, confirmed by an increased module score calculated from the 187 top markers of the CD4 CTL cluster from the 5ʹ PREDICT dataset (**Fig. S4A-C**). The frequency of CD4 CTLs in the two cohorts was higher in IBD patients than in healthy donors (**Fig. S4D-E**), corroborating the findings from our cohort. Moreover, in the adult cohort, which included multiple intestinal regions, CD4 CTLs were enriched across all gut regions of CD patients but only in the affected gut regions (colon, rectum) of UC patients, recapitulating the disease-specific spatial distribution observed in our study (**Fig. S4F**).

We next evaluated whether CD4 CTL frequencies were associated with treatment outcome in the PREDICT study. In both our identification and validation cohorts, pediatric CD patients with partial response to anti-TNF therapy exhibited the highest pre-treatment frequencies of CD4 CTLs in gut biopsies (**Fig. 2I**). In a subset of patients sampled again 3–12 months after initiation of anti-TNF treatment, CD4 CTL frequencies decreased in full responders, whereas in the small number of partial responders analyzed, frequencies did not show an obvious decline (**Fig. 2J**). These findings raise the possibility that elevated CD4 CTL levels at diagnosis may be associated with reduced likelihood of benefit from anti-TNF therapy. In the adult cohort from Thomas et al., we do not observe significant differences in CD4 CTL frequencies before and after treatment with anti-TNF, potentially indicating CD4 CTLs as an arm of the immune response that is not adequately inhibited through anti-TNF blockade in persistent disease (**Fig. S4G**).

Recent work by Rios Martini and colleagues^20^ demonstrated that stimulation of CD4⁺ T cells from adult CD patients with commensal- and food-derived yeasts induces a cytotoxic transcriptional program. We hypothesized that this induced phenotype might resemble the CD4 CTLs we observed in pediatric patients. Indeed, a reanalysis of their *ex vivo* data revealed that yeast-stimulated CD4⁺ T cells from CD patients—but not from healthy donors—acquired a transcriptional profile similar to our CD4 CTL subset, including robust upregulation of cytotoxic effector genes (*GZMB, GZMH, GNLY, IFNG, PRF1*) and characteristic cytokines and receptors (*IL26, IL23R, IL12RB2*) (**Fig. S4H-J**). Out of the three yeast lysates tested, *S. cerevisiae* induced the strongest CD4 CTL phenotype, indicating a potential environmental trigger for CD4 CTL differentiation (**Fig. S4K-L**).

### TCR repertoire analysis reveals clonal expansion of CD4 CTLs

In the 5p PREDICT dataset, we combined gene expression profiling with V(D)J repertoire sequencing to assess the clonality of the CD4 CTL subset. In patients with CD and UC, the most expanded clones were found within CD4 CTLs, Th17 cells, and activated Tregs. In contrast, healthy donors and DGBI patients showed higher clonal expansion of Th2 cells and IELs (**Fig. 3A–B**).

**Figure 3.**
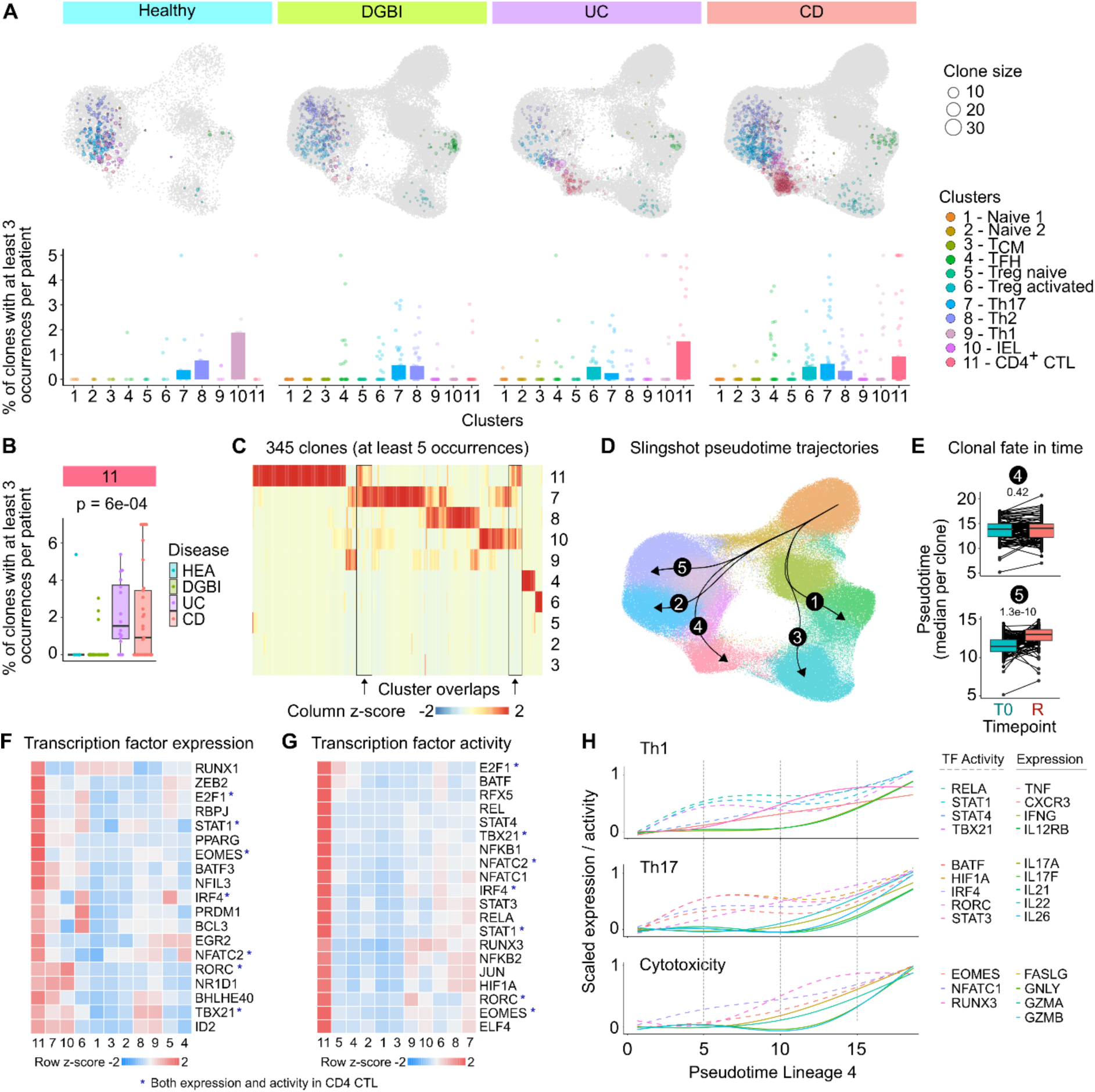
Clonal expansion and transcriptional regulation of CD4 CTL development. **(A)** Clonal expansion of T cells from the 5p gut dataset. Clones were defined as cells sharing identical amino acid sequences of the CDR3β and CDR3α chains. Only clones with at least three occurrences were included. *Top*: Clonal expansion across clusters defined in Fig. 2A. Each dot represents one clone and is positioned at the centroid of the cells belonging to that clonotype; dot size reflects the degree of expansion. *Bottom*: Average clonal expansion across clusters, grouped by participant. Each dot represents one donor. **(B)** Average clonal expansion within the CD4 CTL cluster across participants with DGBI, CD, UC, and healthy donors. Boxplots show the median (center line), interquartile range (box), and whiskers extending to 1.5× the interquartile range. P values were determined using the Kruskal–Wallis test. **(C)** Heatmap showing distribution of cells from individual clonotypes across clusters. Each column represents one clone. Colors indicate column-scaled z-scores of the relative abundance of cells from each clone in each cluster. Black rectangles highlight CD4 CTL clones with mixed cluster identity. **(D)** UMAP projection as in Fig. 2A, colored by pseudotime values calculated using Slingshot. Arrows indicate inferred lineage trajectories. **(E)** Boxplots showing the median pseudotime per clone grouped by time point of sampling in pseudotime lineages 4 and 5. Other lineages are shown in Fig. S6E. T0 – at diagnosis; R – 3 months after diagnosis. Boxplots show the median (center line), interquartile range (box), and whiskers extending to 1.5× the interquartile range. P value was calculated using a two-tailed paired Wilcoxon signed-rank test. **(F)** Heatmap showing relative expression of transcriptional factors (TF) with significantly upregulated expression in cluster 11 compared to other clusters shown in Fig. 2A. Colors represent row-scaled z-scores of average gene expression within each cluster. Rows and cols are hierarchically clustered. **(G)** Heatmap showing the top 20 TFs with the highest transcriptional activity in cluster 11 inferred from gene expression using the CollecTRI transcriptional regulon database. Colors represent row-scaled z-scores of average transcriptional activity within each cluster. Rows and cols are hierarchically clustered. **(H)** Smoothed expression trajectories of selected genes across pseudotime. Gene expression was averaged within pseudotime deciles and normalized to each gene’s maximum value. Lines indicate generalized additive model fits with 95% confidence intervals. Solid lines represent gene expression, and dashed lines represent inferred transcriptional activity based on CollecTRI.

We next examined whether cells sharing the same TCR clonotype were confined to a single transcriptional cluster or distributed across multiple CD4⁺ T cell subsets. Most expanded CD4 CTL clonotypes were composed almost exclusively of cells mapping to the CD4 CTL cluster, indicating limited cross-cluster overlap. In a subset of cases, however, cells from the same clonotype were also detected in Th17 or IEL clusters, suggesting a potential developmental relationship between these states (**Fig. 3C**). Analysis of clone distribution across gut regions showed that identical clones were detected in the duodenum, ileum, and colon (**Fig. S5A–B**). We did not observe substantial clonal sharing between patients, with the exception of two clones each shared across five participants (**Fig. S5C**).

To further investigate developmental trajectories leading to the cytotoxic phenotype, we performed pseudotime analysis using Slingshot^31^ with naïve CD4⁺ T cells specified as the root. This analysis identified five distinct lineages: lineage 1 corresponded to Tfh cells, lineages 2 and 5 to Th1/2 and Th17 cells, lineage 3 to Tregs, and lineage 4 to CD4 CTLs (**Fig. 3D**). Across all lineages, higher pseudotime values were associated with increased clonal expansion, supporting the biological relevance of the inferred trajectories (**Fig. S6A**). To formally test differences in trajectory progression and fate selection, we applied the condiments framework^32^, which confirmed significant differences in fate choices between IBD and non-IBD CD4^+^ T cells (**Fig. S6B–D**).

Because our dataset included repeated sampling from the same participants, we next tracked the fate of individual clones over time. While Tfh, Treg, and CD4 CTL clones remained largely stable between timepoints suggestive of terminal differentiation, Th1/2 and Th17 clones progressed along their respective pseudotime trajectories from the first to the second timepoint (**Fig. 3E**, **Fig. S6E**).

Finally, given suggestions that viral exposure may precede IBD in some individuals^33^, we analyzed TCR sequence similarity to clones previously reported to recognize SARS-CoV-2, CMV, and EBV. Although such clones were detected, they were not enriched in any patient group or within any specific T-cell subset (**Fig. S6F-G**).

As our reanalysis of the Rios Martini et al. dataset^20^ suggested that yeast stimulation can induce a CD4 CTL transcriptional state, we searched for shared TCR clonotypes between the two datasets. Notably, we identified one clonotype which was highly expanded upon *S. cerevisiae* stimulation and which mapped to the CD4 CTL cluster in two CD patients from our cohort (**Fig. S6H**), suggesting a potential link between yeast-reactive responses and the CD4 CTL state in some participants *in vivo*.

### Development of CD4 CTLs is orchestrated by transcription factors

Cytotoxicity in CD4 T cells has been associated with defined transcription factors (TF), including Runx3, Eomes, Tbet (*TBX21*), GATA-3, Bhlhe40, Aiolos (*IKZF3*), and Eos (*IKZF4*)^18,34–38^. We assessed both TF expression and TF activity using downstream target signatures curated in the CollecTRI database^39^. CD4 CTL (cluster 11) displayed high expression of *TBX21, ID2, BHLHE40*, and *RORC*, a pattern shared with Th17 (cluster 7) and IEL (cluster 10) subsets (**Fig. 3F**). In contrast, CD4 CTL uniquely expressed *ZEB2, RUNX1, STAT1, EOMES*, and *RBPJ*, indicating the activation of a transcriptional program distinct from other CD4 lineages.

Although there was partial overlap in TF expression profiles between CD4 CTL, Th17 cells, and IELs, TF activity inferred from target gene modules was entirely distinct in CD4 CTL. The most active TFs inferred in CD4 CTLs included regulators of the Th1 lineage (*STAT1, STAT4, TBX21, RELA*), regulators of the Th17 lineage (*RORC, STAT3, HIF1A, BATF*), and TFs classically associated with CD8 T-cell cytotoxicity (*EOMES, RUNX3*) (**Fig. 3G**). The combined activity of these programs accounted for the defining features of CD4 CTL, including simultaneous expression of Th1 cytokines, Th17 cytokines, and cytotoxic molecules.

Integration of transcription factor activity and gene expression across pseudotime trajectories suggested that CD4 CTLs are transcriptionally related to Th1- and Th17-associated states while additionally engaging a cytotoxic program (**Fig. 3H**). Although these analyses do not establish a definitive developmental path, they support the view that CD4 CTLs represent a distinct effector state combining features of Th1, Th17, and cytotoxic differentiation that are relatively terminally differentiated. Together, the clonality and transcription factor activity data place CD4 CTLs in close relationship to Th1/Th17-associated CD4⁺ T cell programs, while distinguishing them as a separate cytotoxic state.

### Cytokine and cellular interactions associated with CD4 CTL development

Given the unusual transcriptional regulation of CD4 CTL, we next asked which cell–cell interactions and local microenvironmental cues might drive their development. To address this, we integrated scRNA-seq data from ileal LP across three datasets (PREDICT 3p^22^, PREDICT 5p, Thomas et al. 2024^6^), this time including all major lineages rather than restricting the analysis to T cells (**Fig. 4A**, **Fig. S7A–B**). After integration and QC, we obtained a dataset of 1,013,968 high-quality cells spanning nine major lineages: T cells, B cells, endothelial cells, epithelial cells, mast cells, myeloid cells, plasma cells, proliferating cells, and stromal cells, each defined by established marker genes (**Fig. S7C**).

**Figure 4.**
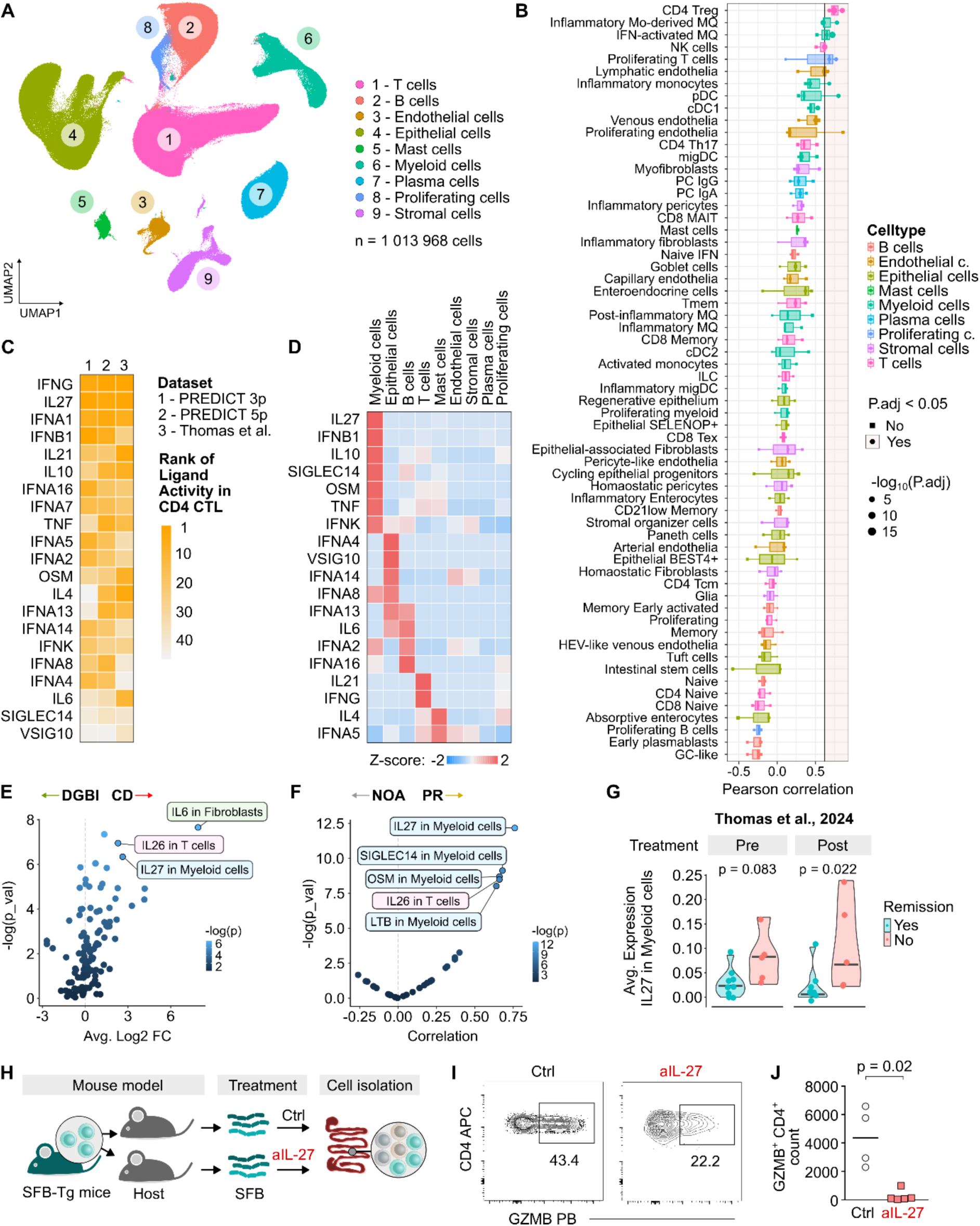
Cell communication networks and functional perturbation identify an IL-27– and interferon-associated cytokine milieu linked to CD4 CTL differentiation. (A–D) Cells from the lamina propria of the ileum from three scRNA-seq datasets (PREDICT 3p, PREDICT 5p, and Thomas et al.) were integrated using STACAS, reclustered, and reannotated (n = 1,013,968 cells from 145 donors). **(A)** UMAP projection of lamina propria cells from the ileum in the integrated dataset, colored by manual annotations. **(B)** Correlations between frequencies of lamina propria cell subsets and frequencies of CD4 CTLs. Correlations were calculated using two-tailed Pearson correlation tests and adjusted for multiple comparisons using the Bonferroni method. Each point represents a correlation from one dataset; point size indicates –log₁₀ adjusted P value. **(C)** Ligand–receptor interactions inferred using NicheNet independently in each dataset. Ligands were scored based on their predicted contribution to CD4 CTL-associated interactions in Crohn’s disease compared with DGBI (PREDICT 3p and 5p) or healthy controls (Thomas et al.). The heatmap shows ranked ligands across datasets. **(D)** Heatmap showing row-scaled z-score of expression of the top-scoring ligands across major lamina propria cell populations shown in (A). **(E–F)** Gene expression in the PREDICT 3p dataset was pseudobulked across major cell populations and participants. **(E)** Volcano plot showing expression of top-scoring ligands in major cell populations from CD versus DGBI samples. Average log₂ fold changes were calculated using Seurat FindMarkers; differential expression was tested using a two-tailed Wilcoxon rank-sum test with Bonferroni adjustment for multiple comparisons. **(F)** Dot plot showing correlations between anti-TNF response (0, NOA; 1, FR; 2, PR) and expression of top-scoring ligands in major cell populations from CD samples. Correlations were calculated using two-tailed Pearson correlation tests with Bonferroni adjustment for multiple comparisons. **(G)** Gene expression in the Thomas et al. dataset was pseudobulked across major cell populations and participants. Violin plot showing average IL27 expression in myeloid cells from CD patients stratified by remission status before and after antiTNF treatment. P values were calculated using a two-tailed Mann–Whitney U test. **(H-J)** SFB-specific T cells were isolated from SFB-Tg mice and adoptively transferred into congenically labeled host mice that were colonized with SFB and treated or not with anti-IL-27 antibody. Intestinal T cells were subsequently isolated and analyzed by flow cytometry. **(H)** Experimental design of the mouse study. **(I)** Representative gating of GZMB⁺ T cells within donor SFB-Tg CD4⁺ T cells. **(J)** Quantification of the absolute counts of GZMB⁺ CD4⁺ T cells. Bars indicate medians. SFB – segmented filamentous bacteria. P value as in G. n = 9 mice in two experiments.

We further subclustered each lineage into fine-grained subsets (**Supplementary Table S2**) and asked which populations showed the strongest correlation in abundance with CD4 CTL across samples and datasets. The top-scoring populations were CD4 Treg, which—similar to CD4 CTL—were markedly expanded in IBD, and two myeloid subsets: inflammatory monocyte-derived macrophages and IFN-activated macrophages (**Fig. 4B**). The prominence of myeloid subsets in this analysis is notable, as these cells are well positioned to provide both antigen presentation and instructive cytokine signals to CD4 CTL. Of note, absorptive enterocytes and intestinal stem cells were negatively correlated with CD4 CTL abundance, suggesting a link between CD4 CTL accumulation and epithelial remodeling in inflamed tissue.

We then applied ligand–receptor interaction analysis using NicheNet^40^ across all three datasets to identify candidate ligands predicted to interact with CD4 CTLs in IBD. Consistently, we observed strong inferred activity of IFN-γ, IL-27, and type I IFN signaling (**Fig. 4C**). *IL27* and type I IFN ligands were predominantly expressed by myeloid subsets, whereas *IFNG* was mainly expressed by T cells (**Fig. 4D**). IFN-γ and type I IFN have been firmly linked to CD pathogenesis^41^, whereas the role of IL-27 is more complex: in CD4 T cells, IL-27 can promote regulatory features but is also a potent driver of cytotoxic differentiation, making it a plausible contributor to the CD4 CTL program^42–47^.

Strikingly, *IL27* expression by myeloid cells was highly enriched in CD compared to DGBI controls and was most pronounced in patients with partial response to anti-TNF treatment (**Fig. 4E–F**). In the Thomas et al. dataset, we observed a similar trend: myeloid *IL27* expression was higher in patients with active disease compared to those in remission (**Fig. 4G**). Together, these data implicate a proinflammatory cytokine circuit involving myeloid-derived IL-27 and type I interferons, together with T cell-derived IFN-γ, as a microenvironmental program associated with CD4 CTL accumulation and treatment failure in CD.

We next tested whether the top candidate ligands from our ligand–receptor analysis, IL-27 and IFN-γ, could induce a cytotoxic phenotype in CD4⁺ T cells activated *in vitro* with CD3/CD28 stimulation to mimic strong TCR signaling (**Fig. S8A-B**). Flow cytometric analysis at 70 h revealed an increase in GZMB, IFN-γ and Tbet, but not GZMA, in CD4⁺ T cells cultured with IL-27 or a combination of IL-27 and IFN-γ (**Fig. S8C**). By contrast, IFN-γ alone did not induce these markers to a similar extent, highlighting a dominant role for IL-27 in promoting this program and supporting the recent findings that CD4 CTLs can be differentiated under specific polarizing conditions *in vitro*^48^.

To test whether IL-27 contributes to CD4 CTL formation *in vivo*, we used a murine adoptive transfer model of CD4 T cells recognizing antigen from segmented filamentous bacteria (SFB)^49^ (**Fig. 4H**). SFB is a murine ileal commensal that induces robust Th17 responses^50^ and promotes conversion of CD4^+^ T cells to GZMB-expressing gut-resident IELs^11^. Four weeks after transfer, SFB-specific donor CD4⁺ T cells upregulated GZMB to levels comparable to CD8⁺ T cells (**Fig. S8D-F**). Blocking IL-27 significantly reduced GZMB expression in SFB-specific CD4⁺ T cells (**Fig. 4I-J**, **Fig. S8G**), supporting a role of IL-27 in acquisition of a cytotoxic program by antigen-specific CD4⁺ T cells *in vivo*.

### Atlas of peripheral blood T and NK cells in IBD and healthy donors

To date, many of the cell subsets identified by scRNA-seq in the diseased intestine are localized to the region of pathology. To overcome the challenges posed by invasive gut biopsies, especially in pediatric settings, we sought to determine whether CD4 CTLs are present in peripheral blood. We analyzed NK and T cells from PBMCs collected at the diagnostic visit prior to treatment initiation, using both 3p and 5p scRNA-seq chemistries, similar to our gut atlas. The 3p dataset included 324,539 cells from 36 donors (**Fig. 5A**, **Fig. S9A**) and the 5p dataset included 593,574 cells from 48 donors (**Fig. 5B**).

**Figure 5.**
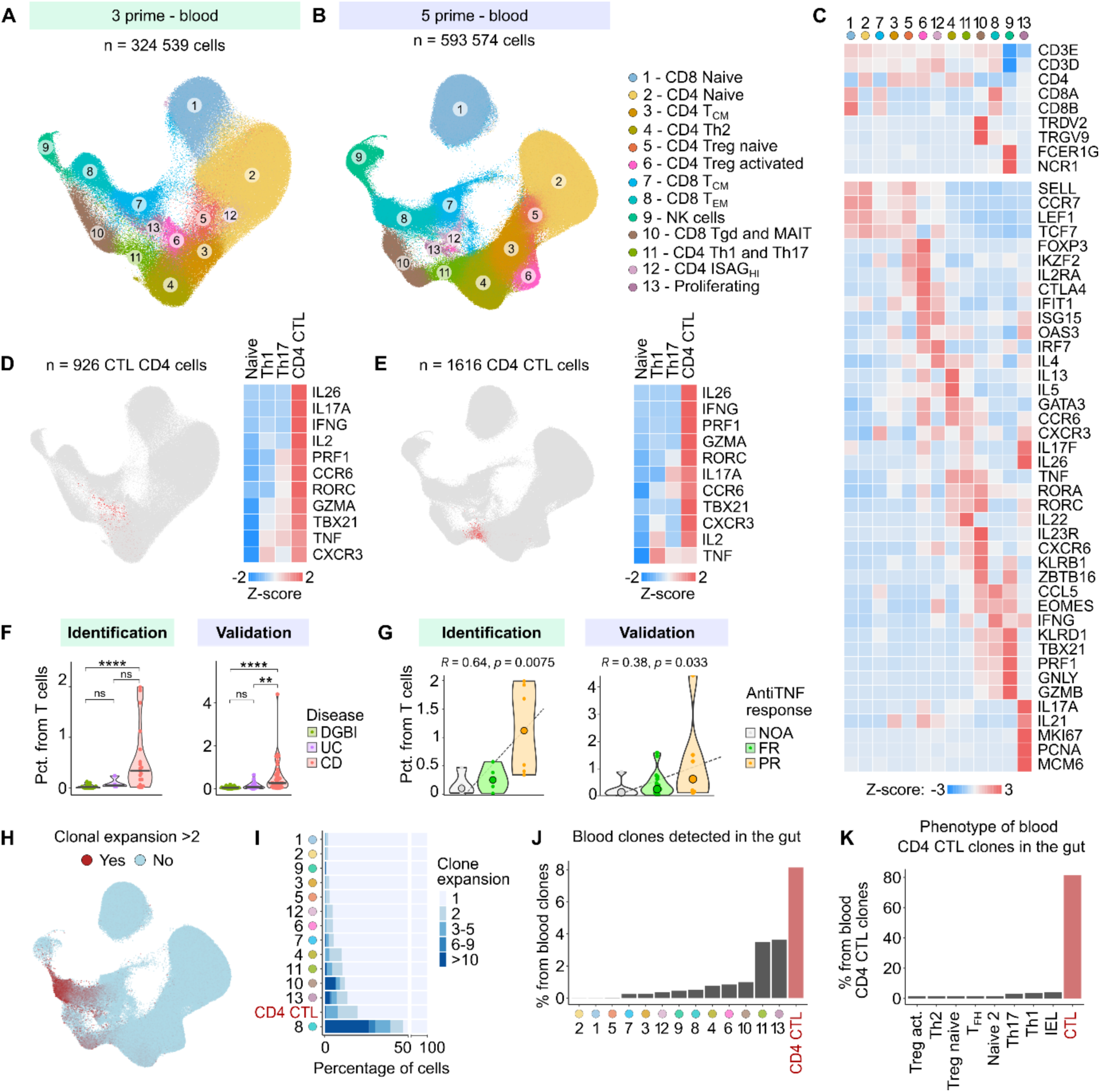
An atlas of peripheral blood T cells in IBD reveals circulating CD4 CTLs. **(A)** UMAP projection of T lymphocytes from the PREDICT 3p PBMC dataset (n = 324,539 cells from 36 donors), colored by manual annotations. **(B)** UMAP projection of T lymphocytes from the PREDICT 5p PBMC dataset (n = 593,574 cells from 48 donors) analyzed separately from 3’ PBMC dataset, colored by manual annotations. **(C)** Heatmap of selected marker genes defining clusters from the PREDICT 5p PBMC dataset shown in (B). Colors represent row-scaled z-scores of average gene expression within each cluster. **(D–E)** Characterization of CD4 CTLs in the PREDICT 3p PBMC dataset (D) and the PREDICT 5p PBMC dataset (E). For details on identification of CD4 CTL see Fig. S9D-E. *Left*: UMAP projections as in (A–B), with CD4 CTLs highlighted in red (n = 926 and n = 1,616 cells, respectively). *Right*: Heatmaps of selected marker genes comparing naïve, Th1, Th17, and CD4 CTL populations. Colors represent row-scaled z-scores of average gene expression within each cluster. **(F)** Percentage of CD4 CTLs among total T cells in 3p participants with CD, UC, and DGBI from the identification and validation cohorts. P values were calculated using two-tailed Mann–Whitney U tests between indicated groups with Bonferroni correction for multiple comparisons. Bars indicate medians. **(G)** Percentage of CD4 CTLs among total T cells in 5p CD participants from the identification and validation cohorts stratified by response to anti-TNF therapy. NOA, not on anti-TNF; FR, full response; PR, partial response. P values were calculated using Pearson correlation tests. Medians are indicated by large dots. **(H)** Analysis of TCR repertoires within CD4⁺ T-cell populations. *Top*: UMAP projection as in (B), with cells colored by clonal expansion status. Expansion is calculated as the log₂ frequency of identical TCR CDR3α and CDR3β sequence pairs in the dataset. **(I)** Quantification of clonal expansion across clusters. Colors indicate grouped clonal expansion values. **(J)** Bar plot showing the percentage of clones from indicated clusters in the PREDICT 5p PBMC dataset that were also detected in the matched PREDICT 5p gut dataset. **(K)** Phenotypic distribution of clones detected within the CD4 CTL cluster in the PREDICT 5p PBMC dataset across corresponding clusters in the matched PREDICT 5p gut dataset. X-axis annotations indicate gut clusters.

Across both datasets, we identified 13 major subsets, with highly concordant gene expression profiles (**Fig. 5C**, **Fig. S9B**). We annotated these subsets using established marker genes. Naïve CD4⁺ (cluster 2) and naïve CD8⁺ T cells (cluster 1), along with CD4 Tcm (cluster 3) and CD8 Tcm cells (cluster 7), expressed *LEF1*, *TCF7*, *SELL*, and *CCR7*. Naïve cells and activated Tregs (clusters 5 and 6) expressed *FOXP3*, *IKZF2*, *IL2RA*, and *CTLA4*. A subset of naïve-like CD4⁺ T cells with high interferon signaling (ISAG^hi^ cells) expressed *IFIT1*, *ISG15*, and *IRF7*^51^.

Among differentiated CD4⁺ T cell subsets, we identified Th2 cells (cluster 4; *GATA3*, *IL4*, *IL5*) and Th1/17-like cells (cluster 11; *CXCR3*, *IL17F*, *TNF*, *IL22*). CD8 effector memory (Tem) cells (cluster 8) and NK cells (cluster 9) expressed cytotoxic molecules (*PRF1*, *GNLY*, *GZMB*). Unconventional T cells (cluster 10) included γδ T cells (*TRDV2, TRGV9*) and MAIT cells (*IL23R*, *KLRB1*, *ZBTB16*), confirmed by TCR rearrangements (**Fig. S9C**). Finally, proliferating cells (cluster 13), characterized by *MKI67*, *PCNA*, and *MCM6*, also expressed Th17-associated cytokines (*IL17A*, *IL17F*, *IL21*).

### Blood CD4 CTLs are increased in IBD and correlate with anti-TNF response

To improve resolution of CD4 states that may be obscured by joint clustering with CD8⁺ T cells and NK cells, we subset differentiated CD4⁺ T cell clusters and performed a focused reanalysis (**Fig. S9D-F**). In both the 3p and 5p datasets, this approach revealed a distinct cluster of CD4⁺ T cells co-expressing type 1 (*IFNG, TBX21*) and type 17 (*IL17A, RORC*) markers, together with cytotoxic effector genes (*GZMA, PRF1*) (**Fig. 5D-E**). These transcriptional profiles were highly similar to gut CD4 CTL cells (**Fig. S9G-H**) and the frequencies of blood CD4 CTL cells highly correlated with frequencies of CD4 CTL cells in the ileum and colon (**Fig. S9I**).

We next quantified the frequencies of peripheral CD4 CTLs across disease groups. CD4 CTLs were significantly increased in patients with CD compared to DGBI controls (**Fig. 5F**). Notably, in CD, the frequency of circulating CD4 CTLs was associated with clinical non-response to anti-TNF therapy, with the highest frequencies observed in patients exhibiting partial response (**Fig. 5G**).

To investigate clonal relationships, we analyzed TCR repertoires in the 5p dataset. As expected, the highest levels of clonal expansion were observed in CD8 Tem, proliferating cells, and MAIT cells (**Fig. 5H–I**). Remarkably, CD4 CTLs also showed substantial clonal expansion, with expanded clones representing over 20% of this subset (**Fig. 5I**). This prompted us to examine potential overlap between peripheral and gut-resident CD4 CTL clones. Indeed, we identified shared clonotypes between peripheral blood and gut CD4⁺ T cells. Among CD4 CTLs, 8% of peripheral clones were also detected in the gut of the same patients, which represented a value double that even seen for Th1 and Th17 cells (cluster 11) and proliferating cells (cluster 13) (**Fig. 5J**). Notably, the matched gut cells almost exclusively exhibited the CD4 CTL phenotype (**Fig. 5K**).

Together, these findings demonstrate that CD4 CTLs are not only expanded in the inflamed gut but are also detectable and clonally related in the peripheral blood of CD patients, supporting their potential as noninvasive recirculating biomarkers of disease activity and treatment response.

### Detection of CD4 CTLs by flow cytometry

Next, we aimed to determine whether peripheral blood CD4 CTLs could be detected using flow cytometry. Based on their transcriptional profile, we hypothesized that CD4 CTLs could be phenotypically defined as CD3⁺ CD4⁺ CD8⁻ CXCR6⁺ CD27⁻ cells (**Fig. 6A**, **Fig. S10A**). In PBMC samples from the PREDICT cohort, such gating identified cells which were virtually absent in blood from DGBI patients and healthy donors but were significantly increased in both UC and CD patients with the average frequency of 0.2-0.3% from CD3+ T cells (**Fig. 6B**). To further characterize their cytotoxic potential, we stained for GZMA, GZMB, PRF1, and the transcription factor T-bet. These cytotoxic markers were significantly upregulated in CD4⁺ CXCR6⁺ CD27⁻ cells compared to CD27^+^ naïve CD4⁺ T cells (**Fig. 6C**, **Fig. S10B**). Notably, T-bet expression in CD4⁺ CXCR6⁺ CD27⁻ cells was comparable to that observed in CD8⁺ T cells, while GZMA levels even exceeded those in CD8⁺ T cells.

**Figure 6.**
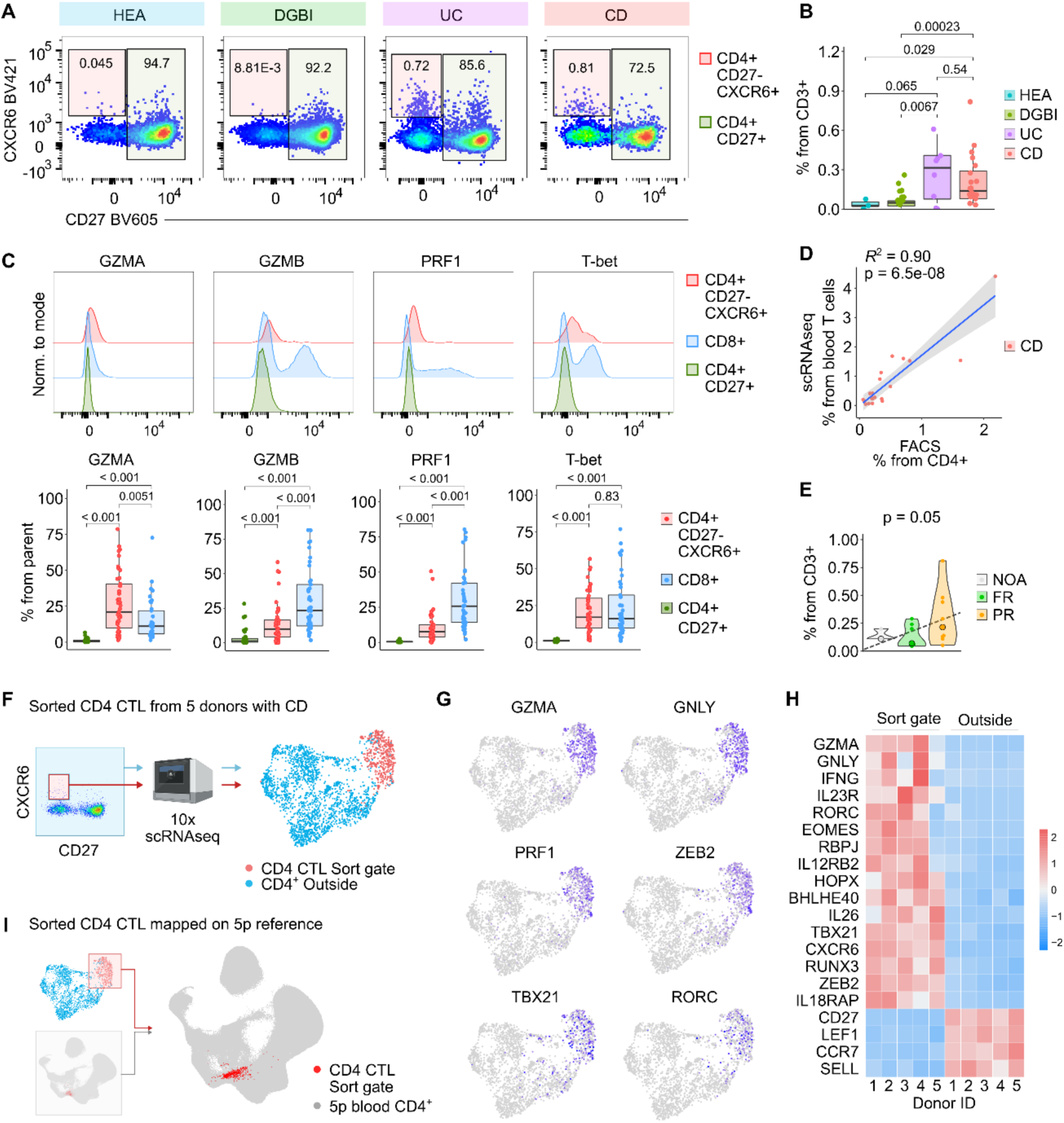
CD4⁺ CXCR6⁺ CD27⁻ gating enables functional and transcriptional validation of circulating CD4 CTLs. **(A)** Gating strategy for identification of CD4 CTL cells by flow cytometry, defined as CD4⁺ CD27⁻ CXCR6⁺ T cells. Representative plots show CD4 CTL cells in a healthy donor and donors with DGBI, ulcerative colitis (UC), and Crohn’s disease (CD). The full gating strategy is provided in Fig. S10. **(B)** Proportions of CD4 CTL cells (CD4⁺ CD27⁻ CXCR6⁺) among CD3⁺ T cells in healthy donors and donors with DGBI, CD, and UC from the PREDICT cohort. P values were calculated using two-tailed Mann–Whitney U tests with Bonferroni correction for multiple comparisons. Boxplots show the median (center line), interquartile range (box), and whiskers extending to 1.5× the interquartile range. **(C)** Representative histograms (top panels) and quantification of the percentage of positive cells (bottom panels) for GZMA, GZMB, PRF1, and T-bet in CD4 CTL cells (CD4⁺ CD27⁻ CXCR6⁺), CD4⁺ CD27⁺ T cells, and CD8⁺ T cells. The full gating strategy is provided in Fig. S10. P values and boxplot definitions are as in (B). **(D)** Correlation between the frequency of CD4 CTL cells measured by flow cytometry and detected by scRNA-seq in peripheral blood samples from the same CD participants. P value was calculated using a two-tailed Pearson correlation test. **(E)** Percentage of CD4 CTL cells in peripheral blood of CD patients grouped by response to anti-TNF therapy. NOA – not on anti-TNF therapy; FR – full response; PR – partial response. P value was calculated using one-sided Pearson correlation test. **(F)** Schematic illustration of sorting CD4 CTL cells for scRNA-seq validation (left) and UMAP projection of sorted CD4 CTL cells and control CD4 T cells (right). n = 2,510 cells from 5 donors. **(G)** UMAP projection from (F) showing the expression of GZMA, GNLY, PRF1, ZEB2, TBX21, and RORC. **(H)** Heatmap showing the expression of selected marker genes in CD4 CTL cells and control CD4⁺ non-CTL cells across the 5 donors. Colors represent row-scaled z-scores of average gene expression. **(I)** Sorted CD4 CTL cells from scRNA-seq were mapped onto the 5p peripheral blood dataset (UMAP projection from Fig. 5B) using Azimuth.

To further confirm that these flow cytometry–defined cells correspond to the CD4 CTLs identified by scRNA-seq, we analyzed a subset of matching samples profiled by both methods. We observed a strong correlation between the frequency of CD4 CTLs detected by scRNA-seq and the frequency of CD4⁺ CXCR6⁺ CD27⁻ T cells measured by flow cytometry (**Fig. 6D**), confirming that this gating strategy may accurately capture the CD4 CTL population. The frequencies of CD4⁺ CXCR6⁺ CD27⁻ T cells were highest in patients with partial response to anti-TNF therapy (**Fig. 6E**), highlighting the potential of this flow cytometry-based approach to capture a feature associated with anti-TNF treatment resistance.

### Independent validation of CD4 CTL identity by targeted scRNA-seq

To independently validate the transcriptional phenotype of CD4 CTLs identified by flow cytometry, we performed targeted scRNA-seq on sorted cells. Specifically, we isolated CD3⁺ CD4⁺ CD8⁻ CXCR6⁺ CD27⁻ T cells from the peripheral blood of five new donors with severe, treatment-refractory CD who were not part of any previous cohort (**Supplementary Table S3**). As controls, we also sorted non-CTL CD4⁺ T cells (i.e., CD4⁺ T cells excluding the CXCR6⁺ CD27⁻ subset) from the same samples (**Fig. 6F**, **Fig. S10C**).

Compared to the control CD4⁺ T cells, sorted CD4⁺ CXCR6⁺ CD27⁻ cells showed markedly higher expression of cytotoxic molecules, including *GZMA*, *GNLY*, and *PRF1* (**Fig. 6G-H**), a pattern consistently observed across donors (**Fig. 6H**). To assess transcriptional similarity, we mapped the sorted cells to our 5p reference atlas using Azimuth^52^. This analysis confirmed that the sorted population aligned closely with the CD4 CTL cluster previously identified in our scRNA-seq dataset (**Fig. 6I**), further supporting the robustness and reproducibility of this cell state across technologies and patient cohorts.

## Discussion

The immunopathologic mechanisms driving treatment resistance in IBD remain poorly understood, and current therapies such as anti-TNF biologics benefit only a subset of patients. To address this, we performed high-resolution single-cell transcriptomic profiling of T cells from treatment-naïve pediatric IBD patients.

We revealed a transcriptionally unique subset of CD4+ T cells with cytotoxic phenotype marked by expression of *GZMA, GZMB, GNLY, PRF1*, and *IFNG*, alongside Th1- and Th17-associated transcripts including *TBX21, CXCR3, CXCR6, RORC, IL23R,* and *IL26*. These cells were clonally expanded, suggesting antigen-driven activation, and were enriched in inflamed intestinal tissue from IBD patients. Their localization mirrored disease distribution (ileum in CD, colon in UC), and in CD, their frequency was highest in patients who subsequently showed poor response to anti-TNF therapy. Importantly, we also identified the CD4 CTL subset within peripheral blood.

While several studies have recently reported on the T cell subsets enriched in CD and UC, a clear CD4 CTL subset has not been defined in these studies^4–6,21^. However, when we reanalyzed two studies, we found CD4 CTLs which were highly transcriptionally matched to those identified in the PREDICT cohort. The key analytical step we took was to computationally isolate CD4 T cells away from NK cells and CD8 CTLs which influence the relative enrichment of cytotoxic genes as markers for CD4 CTLs in this mixed analysis. CD4 CTLs have been reported in multiple settings including chronic viral infections, cancer, and aging^37,53–60^. However, in IBD their origin, functional relevance, and relationship to clinical outcomes remain incompletely defined. Our findings extend this emerging literature by defining a clonally expanded CD4 CTL state in treatment-naïve pediatric IBD and linking its abundance to subsequent anti-TNF response.

CD4⁺ T cells play a central role in mucosal immunity and IBD pathogenesis. In inflamed tissue, sustained exposure to microbial- and dietary-derived antigens presented on MHC class II by epithelial and antigen-presenting cells is thought to contribute to ongoing CD4⁺ T cell activation^11,12,61^. The CD4 CTLs identified here combined inflammatory cytokine expression with cytotoxic effector molecules, suggesting potential contributions to epithelial injury and chronic inflammation. Notably, major therapeutic classes used in IBD—including anti-TNF agents, anti-integrin therapy, anti-IL-12/23, and JAK inhibitors—target pathways that intersect with these effector programs^62,63^. Whether and how these therapies modulate CD4 CTL abundance or function in the gut remains an important question.

The differentiation of effector CD4⁺ T cell subsets is shaped by signals from the local tissue microenvironment. Through cell–cell interaction analysis of our scRNA-seq datasets from inflamed ileal tissue, we found that CD4 CTL development is associated with exposure to type I and type II interferons, as well as IL-27—cytokines known to promote cytotoxic T cell programs^64–66^. Type I interferons and IL-27 were predominantly expressed by myeloid cells, suggesting that these cells may serve as both antigen-presenting and cytokine-producing drivers of CD4 CTL differentiation. In contrast, IFN-γ was produced primarily by CD4 CTLs themselves, potentially reinforcing a self-amplifying inflammatory loop within the tissue.

While these transcriptional interactions suggest a cytokine-driven circuit for CD4 CTL development, their spatial context remains unknown. Recent spatial transcriptomics atlases of CD and UC have revealed compartmentalized immune cell niches and altered cellular interactions in inflamed gut tissue^67,68^. Future spatially resolved studies will be essential to determine the localization of CD4 CTLs within the mucosa, their proximity to antigen-presenting and cytokine-producing cells, and how these interactions evolve with therapy.

An important unanswered question is which antigens are recognized by CD4 CTLs in IBD. Addressing antigen specificity is challenging because CD4 CTLs are infrequent, current computational methods cannot reliably infer specificity from TCR sequence alone, and HLA heterogeneity complicates both *in silico* prediction and *ex vivo* testing. Nonetheless, recent work has reported enhanced CD4⁺ T cell reactivity to yeast antigens in CD, which was associated with acquisition of cytotoxic phenotype *ex vivo*^20^. Consistent with this, we identified a TCR clonotype shared between yeast-stimulated CD4⁺ T cells in that dataset and CD4 CTLs from two CD patients in our cohort, where it mapped specifically to the CD4 CTL cluster. Although anecdotal, this observation raises the possibility that some CD4 CTLs recognize *S. cerevisiae*–derived antigens and supports the broader hypothesis that persistent microbial antigen exposure contributes to CD4 CTL differentiation in the inflamed gut.

Finally, we demonstrate that CD4 CTLs are not restricted to the intestinal mucosa in pediatric CD but are also detectable in peripheral blood by both scRNA-seq and flow cytometry. This supports the feasibility of measuring circulating CD4 CTLs using a clinically accessible assay and identifies them as a candidate blood-based biomarker for baseline treatment stratification. Furthermore, it may also provide a route for them to recirculate and be recruited to distal regions of the intestine beyond the initial site of inflammation.

In summary, our study defines an IBD-associated cytotoxic CD4⁺ T cell state that is clonally expanded, enriched at disease-relevant intestinal sites, and linked to poor response to anti-TNF therapy. Prospective validation in larger and independent cohorts will be required to establish whether quantification of CD4 CTLs at diagnosis can support patient stratification, guide therapeutic decision-making in IBD, and whether they present a subset which is causal and targetable for therapy.

## Methods

### Participant recruitment and diagnosis

Participant enrollment in the study was described previously^22,23^. The PREDICT trial (ClinicalTrials.gov, #NCT03369353) enrolled pediatric participants aged 6 years or older and weighing at least 10 kg. Enrollment took place at Fred Hutchinson Cancer Center (Seattle, Washington, USA) or Boston Children’s Hospital (Boston, Massachusetts, USA) between May 1, 2017, and November 25, 2024.

Ethical protocols were approved by the Fred Hutchinson Cancer Center Institutional Review Board (Protocol #9730), the Boston Children’s Hospital Institutional Review Board (Protocol #IRB-P00030890), and the Seattle Children’s Institutional Review Board (Protocol #1638). Written informed consent was obtained, with assent when applicable.

Diagnoses of DGBI, CD, and UC were made by the patient’s clinician based on endoscopic, histopathologic, and clinical features. CD patients were followed prospectively and categorized as FR, PR, or NOA. FR to anti-TNF was defined as clinical symptom control and biochemical response, with a wPCDAI score <12.5 on maintenance anti-TNF therapy. PR was defined as lack of clinical symptom control and biochemical response, with documented escalation of anti-TNF therapy at 2 years after the baseline visit.

Healthy donors were recruited through two protocols. Endoscopy participants were recruited at Seattle Children’s Hospital for collection of blood, stool, urine, and gastrointestinal tissue during research endoscopy under Seattle Children’s Institutional Review Board Protocol #1638. Eligibility criteria included

age 18–25 years, no acute or chronic illness, no chronic medications other than vitamin supplements or birth control, body mass index (BMI) of 20–25, and not being pregnant. Additional healthy donors were recruited through the PREDICT study at Boston Children’s Hospital for peripheral blood collection under Boston Children’s Hospital Institutional Review Board Protocol #IRB-P00030890. Eligibility criteria included age 18 years or older, no acute illness, no known immune-related disease, and no use of immunosuppressive medications. Recruitment used flyers and social media advertisements.

### Collection and processing of peripheral blood

Whole blood was drawn into EDTA anticoagulant. 2mL of whole blood were aliquoted into 2x 1mL vials and were frozen at −80° C. The remaining blood was used for isolation of PBMCs via the standard method using Ficoll- Paque Plus density gradient media. Isolated PBMCs were then cryopreserved at ∼5 × 10^6^ cells/vial with 90% FBS/10% DMSO freezing medium. Cells were slow-frozen using a Mr. Frosty container at -80C for 24hrs and then transferred to -150C for long term storage.

### Collection and processing of intestinal biopsies

Intestinal biopsies were processed as described previously^22,23^. Pinch biopsies were taken from the duodenum, terminal ileum, and colon, prioritizing areas with marked erythema or edema; left-sided biopsies were obtained by default. Biopsies were placed in RPMI 1640 (Thermo Fisher Scientific, #21870-076) on ice. All centrifugation steps were performed at 4°C.

Biopsies were rinsed in 25 mL PBS (Thermo Fisher Scientific, #14190-144) and transferred to 10 mL ECS (HBSS, Ca/Mg-free, (Thermo Fisher Scientific, #14175-103), 100 U/mL penicillin + 100 μg/mL streptomycin (Thermo Fisher Scientific, #15140-122), 10 mM HEPES (Thermo Fisher Scientific, #15630-080), and 2% FBS (Thermo Fisher Scientific, #SH3007103HI)) freshly supplemented with 10 mM EDTA (Thermo Fisher Scientific, #AM9261). EPI and LP were separated for 15 min at 37°C with rotation (700 rpm), followed by 10 min on ice, vigorous shaking (20×), and high-speed vortexing (10 s).

The LP tissue fraction was carefully removed and rinsed in 45 mL ice-cold PBS and digested in 10 mL enzymatic mix (RPMI 1640 (Thermo Fisher Scientific, #21870-076), penicillin (Thermo Fisher Scientific, #15140-122), streptomycin (Thermo Fisher Scientific, #15140-122), 10 mM HEPES (Thermo Fisher Scientific, #15630-080), 2% FBS (Thermo Fisher Scientific, #SH3007103HI), and 50 μg/mL gentamicin (Thermo Fisher Scientific, #15750-060)) supplemented with 100 μg/mL Liberase TM (Roche, #5401127001) and 100 μg/mL DNase I (Sigma, #D5025-150KU) for 30 min at 37°C with rotation (700 rpm). Dissociation was quenched with 80 μL 0.5 M EDTA, incubated on ice for 5 min, triturated, filtered through a 40 μm strainer (VWR, #21008-949), and brought to 35 mL with ice-cold PBS. Cells were centrifuged at 500g for 10 min, resuspended in 1.2 mL ECS, centrifuged at 800g for 2 min, treated with ACK lysis buffer (Gibco, #A10492-01) for 3 min on ice, quenched with 500 μL 0.2% FBS (Thermo Fisher Scientific, #SH3007103HI) in DPBS (Thermo Fisher Scientific, #14190-144), centrifuged at 800g for 2 min, resuspended in PBS with 0.4% BSA (Thermo Fisher Scientific, #AM2616), filtered, washed, centrifuged at 500g for 5 min, and resuspended in 60 μL PBS with 0.4% BSA.

For EPI, ECS was added to 35 mL, centrifuged at 600g for 10 min, and resuspended in 1 mL ECS. Cells were pelleted twice (800g for 3 min) in a 0.4% BSA-coated tube, treated with ACK lysis buffer (Thermo Fisher Scientific, #A10492-01) for 2 min on ice, washed in ECS without FBS, and dissociated in TrypLE Express (Thermo Fisher Scientific, #12604-013) (1 mL + 400 μL) for 4 min at 37°C with trituration, followed by 3 min + 3 min at 37°C with trituration in between. Cells were centrifuged at 800g for 3 min, resuspended in PBS with 0.4% BSA, filtered through a 40 μm strainer, washed, centrifuged at 500g for 5 min, and resuspended in 50 μL PBS with 0.4% BSA.

Cells from EPI and LP fractions were counted and prepared as single-cell suspensions for scRNA-seq.

### ScRNA-seq – 3ʹ protocol

Single cells were loaded onto 3ʹ library chips according to the manufacturer’s protocol for Chromium Single Cell 3ʹ Library (v2) (10x Genomics, #PN-120237, #PN-120236, #PN-120262). Cells were partitioned into GEMs for barcoded reverse transcription, followed by cDNA amplification, enzymatic fragmentation, and 3ʹ adapter and sample index attachment. Libraries were sequenced on HiSeq or NovaSeq instruments (read 1, 26 bp; read 2, 91 bp; index 1 (i7), 8 bp), and quality-filtered base calls were converted to demultiplexed FASTQ files. FASTQ files were aligned to GRCh38 using Cell Ranger (v2.2) on the Cumulus/Terra pipeline to generate cell-by-gene matrices per sample. Matrices were imported into Seurat and annotated using SingleR (v1.0) with the Human Primary Cell Atlas reference^69,70^. T and NK cells were retained for subsequent analyses.

### ScRNA-seq – 5ʹ protocol

Single cells were loaded onto 5ʹ library chips according to the manufacturer’s protocol for Chromium Single Cell 5ʹ Library and Gel Bead Kit (10x Genomics). Biopsies were processed using 5ʹ v1 kits (prior to November 2020) (10x Genomics, #PN-1000006; #PN-1000005) or 5ʹ v2 kits (since November 2020) (10x Genomics, #PN-1000263; #PN-1000252). For libraries prepared using the 5’ v2 assay sequencing was performed on Illumina NovaSeq 6000 (GEX libraries: Read 1 26-bp, Read 2 80-bp, 10-bp i7 and 10-bp i5 reads; V(D)J libraries: Read 1 150-bp, 10-bp i7, 10-bp i5, Read 2 150-bp). For libraries prepared using the 5’ v1 assay, sequencing was performed on Illumina NovaSeq 6000 (GEX libraries: Read 1 26-bp, Read 2 80-bp, 8-bp i7 and 0-bp i5 reads; V(D)J libraries: Read 1 150-bp, 8-bp i7, 0-bp i5, Read 2 150-bp).

FASTQ files were aligned to GRCh38 reference genome using Cell Ranger (v5.0.1) to generate cell-by-gene matrices per sample. V(D)J reads were aligned using Cell Ranger (v5.0.1) against the GRCh38-alts-ensembl-2.0.0 reference, and filtered_contig_annotations were joined to the Seurat object using a custom function^71^. For the clonal expansion analyses, cells with the same amino acid sequences of the CDR3α and CDR3β were considered clones.

Initial preprocessing was performed separately for PBMC and tissue samples. Cell-level QC considered library complexity, UMI counts, detected features, and the proportion of mitochondrial, ribosomal, hemoglobin, and viral transcripts. Cells with >30% mitochondrial reads, <250 UMIs, or <100 detected genes were excluded. Genetic demultiplexing was used to identify cross-donor doublets, and Scrublet was used to support identification of heterotypic doublets. Gene counts were normalized and log-transformed using Scanpy, highly variable genes were identified, dimensionality reduction was performed by PCA, and batch effects were corrected using Harmony. Broad cell type annotation was performed iteratively based on canonical marker genes, with Celltypist, SCimilarity, and ARBOL used as supportive annotation tools.

For the present study, T/NK cells and proliferating cells were retained for downstream analyses. For each dataset, samples were merged, re-normalized, scaled, and analyzed by PCA (20 PCs), UMAP, and Louvain clustering; low-quality clusters were removed and processing was reiterated. Final datasets of 5ʹ T/NK and proliferating cells were integrated by donor ID using STACAS (v2.2.2).

### Analysis and visualization of scRNA-seq data

Transcriptional regulatory interactions were estimated using the decoupleR package (v2.10.0) using the CollecTRI database^39^. GSEA pathways were processed using the R package fgsea (v1.20.0)^72^. Heatmaps were created with the R package pheatmap (v 1.0.12). DEGs were identified using the two-sided Wilcoxon rank-sum test (Seurat FindMarkers), with p-values adjusted for multiple testing using the Bonferroni correction.

Pseudotime analysis was performed using the *slingshot* package (v2.12.0)^31^ to infer a lineage trajectory among gut T-cell subsets based on UMAP embeddings and cluster annotations. To test for differences in pseudotime progression between groups, we used the *condiments* package (v1.12.0)^32^, which estimates group-specific changes in pseudotime distributions along the inferred trajectory.

Ligand-receptor interaction analyses were performed using *NicheNet* (v2.2.0)^40^. Analyses were conducted independently for three scRNA-seq datasets: PREDICT 3p, PREDICT 5p, and the Thomas et al. dataset. In each dataset, CD4 CTL cells were defined as receiver cells. Sender cell populations comprised all remaining annotated cell types. Differential expression analysis was performed within all T cells to define condition-specific gene expression signatures, comparing DGBI versus CD in PREDICT 3p and PREDICT 5p, and Healthy versus CD in the Thomas et al. dataset. DEGs were used as input gene sets to infer upstream ligands. For each sender cell type, expressed ligands were prioritized based on their regulatory potential scores, which quantify the predicted ability of each ligand to regulate the receiver gene signature through curated ligand–receptor relationships.

### Analysis of previously published datasets

For the Thomas et al. dataset^6^, processed .h5ad files were downloaded from Zenodo (doi:10.5281/zenodo.13768607) and used to create Seurat objects for analysis in R.

For the Elmentaite et al. dataset^21^, gene-by-cell count matrices and metadata were obtained from the Human Cell Atlas Data Portal and used to create Seurat objects for analysis in R. Only T cells were analyzed, subsetted based on author annotations (“Activated CD4 T”, “SELL+ CD4 T”, “Tfh”, “Th1”, “Th17”, “Treg”).

For the Rios-Martini et al. dataset^20^, gene-by-cell count matrices and metadata were obtained from GEO (GSE227638) and used to create Seurat objects for analysis in R. TCR data (filtered_contig_annotations) were obtained from GEO (GSE227638) and merged with Seurat objects using the same custom R function used for PREDICT 5p data.

All datasets were processed using the same pipeline, including normalization, scaling, dimensional reduction (PCA and UMAP), and Louvain clustering. Datasets were integrated by Donor ID using STACAS. CD4 CTL cells were identified among T cells by clustering and manual annotation. Similarity to PREDICT 5p CD4 CTL cells was quantified using AddModuleScore, applying a PREDICT 5p CD4 CTL gene signature to each dataset.

### Flow cytometry

The list of used antibodies is provided in Supplementary Table S4. Flow cytometry was performed in six batches. Cryopreserved human PBMCs were thawed rapidly at 37 °C, diluted dropwise into prewarmed RPMI 1640 (Thermo Fisher Scientific, #11875119) supplemented with 10% FBS (Gemini, # S11550) and antibiotics (Thermo Fihser Scientific, # 15140-22), and washed by centrifugation (300g, 5 min, RT). Cells were resuspended in complete medium with recombinant human IL-7 (10 U/mL; R&D Systems, #207-IL) and rested for 1–2 h at 37 °C, 5% CO₂ before use.

Cells were stained with Zombie NIR Fixable Viability Kit (BioLegend, #423106), fixed and permeabilized using the Foxp3/Transcription Factor Staining Buffer Set (ThermoFisher, #00-5523-00), and stained with antibodies in Permeabilization buffer supplemented with Brilliant Stain Buffer (ThermoFisher, #00-4409-42). Intracellular staining was performed overnight at 4 °C before acquisition.

### scRNA-seq – validation experiment

Cryopreserved PBMCs from five donors with severe CD (list of donors provided in Supplementary Table S3) were thawed as described above. Cells were labeled with antibodies listed in the Supplementary Table S4. Live CD3+ CD4+ CD8- CXCR6+ CD27- cells were sorted (total count of sorted cells 1742) and other Live CD3+ CD4+ CD8- cells were taken as control (2000 cells from each donor, 10000 cells in total). Cells were loaded on 10x Chromium and processed with Chromium GEM-X Single Cell 5’ v3 (10x Genomics, # PN-1000699, PN-1000694, PN-1000698, PN-1000252, PN-1000215). Libraries were sequenced on a NovaSeq X 25B 100 cycle kit with the 10X recommended read structure I1/I2/R1/R2 10/10/28/90. Donor identities were demultiplexed using popscle Freemuxlet (v1)^73^.

### In vitro stimulation of human PBMCs

Cryopreserved PBMCs from donors with CD, UC, DGBI and healthy donors from the PREDICT cohort were thawed as described above. For proliferation assays, cells were labeled with CellTrace Violet (ThermoFisher, #C34571) according to the manufacturer’s instructions. For T cell activation, PBMCs were seeded at 1.5 × 10⁵ cells per well in round-bottom 96-well plates (Corning, #3799) and stimulated with Dynabeads™ Human T-Activator CD3/CD28 (ThermoFisher, #11131D) at a 1:1 cell-to-bead ratio in 200 µL of RPMI 1640 (Thermo Fisher Scientific, #11875119) supplemented with 10% FBS (Gemini, # S11550) and antibiotics (Thermo Fihser Scientific, # 15140-22). In some experiments, 100 ng/mL recombinant human IFNG (R&D systems, #285-IF-100) and/or 100 ng/mL recombinant human IL-27 (R&D systems, # 2526-IL-010) were added to the culture. Cells were cultured for 70 h at 37 °C, 5% CO₂. Protein transport inhibitor Brefeldin A, (BioLegend, #420601) was added for the final 4 h of stimulation. At the end of culture, cells were harvested, beads were removed using a magnetic stand, and samples were prepared for flow cytometry. Cells were stained as described above.

### Adoptive transfer of SFB-specific CD4+ T cells

Peripheral (axillary, brachial, and inguinal) and mesenteric lymph nodes were isolated from donor SFBtg (CD90.1/2) or WT (CD90.1) mice^49^ and processed into single-cell suspensions. Cells were stained with antibodies against TCRβ, CD4, CD8α, CD8β, and CD62L and sorted on a cell sorter (MA900, SONY).

Naïve CD4⁺ SFBtg T cells (TCRβ⁺ CD4⁺ CD62L⁺) were co-transferred with polyclonal WT lymph node cells into Rag1⁻/⁻ recipient mice^74^ by intravenous injection (2 × 10⁴ SFBtg and ∼2 × 10⁵ WT cells per mouse). Where indicated, mice received intraperitoneal injections of anti–IL-27 antibody (250 μg, clone MM27.7B1, Bio X Cell, # BXC-BE0326) or PBS control on the same day as T cell transfer, administered twice within 24 h.

Small intestinal IELs were isolated using a protocol as previously described^11^. Briefly, the small intestine was dissected, mesenteric fat and Peyer’s patches were removed, and equal segments of duodenum, jejunum, and ileum were opened longitudinally, cut into ∼2-cm pieces, and washed in ice-cold PBS. Tissue fragments were incubated twice for 20 min at 37 °C in HBSS supplemented with 3% FBS and 2 mM EDTA with gentle agitation, followed by vigorous vortexing to release epithelial cells and associated IELs. Tissue remnants were removed, and the resulting cell suspension was filtered through a 100-μm cell strainer, pelleted by centrifugation (300 g, 8 min, 4 °C), and stained for flow cytometric analysis. The list of used antibodies is provided in Supplementary Table S5.

### Statistical analysis

Statistical analyses were performed in R v4.2.1 using the tests specified in the figure legends. Unless otherwise stated, scRNA-seq and flow cytometry data were analyzed using two-tailed nonparametric Mann–Whitney U tests. Where appropriate, alternative tests were used and are indicated in the corresponding figure legends. Unless otherwise stated, all statistical tests were two-sided. One-sided tests were used only for validation analyses of previously observed directional findings and are indicated in the relevant figure legends. Multiple-comparison correction was applied for differential expression and other exploratory analyses involving multiple testing. By contrast, flow cytometry analyses were performed on predefined marker panels selected for targeted validation, and correction for multiple comparisons was not applied unless explicitly stated. The number of biological replicates (cells, donors, or mice) is provided in the respective figure legends.

## Data availability

The cell-by-gene matrices will be available for download with the peer-reviewed version of this manuscript and are currently viewable in the Broad Single Cell Portal (SCP) under study accession numbers SCP3737 (blood), SCP3739 (total CD3 and NK cells from gut), and SCP3741 (CD4 T cells from gut). The raw human FASTQ files will be available from the Broad controlled access repository DUOS with the peer-reviewed version of this manuscript. Data from previously published studies used for validation of our findings are available in the following databases: datasets from Thomas et al. are available at Zenodo (https://doi.org/10.5281/zenodo.13768607), datasets from Rios-Martini et al. are available at Gene Expression Omnibus (accession no. <u>GSE227638</u>) datasets from Elmentaite et al. are available at the Gut Cell Atlas (https://www.gutcellatlas.org/).

## Code availability

The code can be accessed at https://github.com/vercanie/predict_cd4_ctl/.

## Author Contributions Statement

Conceptualization, VN, AKS, LSK, JO-M; Methodology, VN, KKi, MEB, RH, JV, JD, OS, AKS, LSK, JO-M; Software, VN, KKi, MEB, RH, LN, ZS; Validation, VN, IP, JDSC, JV, KKo; Formal Analysis, VN, KKi, MEB, RH, JV, LN, ZS; Investigation, VN, KKi, HBZ, MEB, RH, PK, IP, JDSC, JV, AK, LN, ZS, JO-M, KKo, BB, JZ, WKL, SM, NF, ZH, GW, DS, LA, DL; Resources, HBZ, MEB, PK, LN, ZS, SBS, JD, OS, AKS, LSK, JO-M, BB, JZ, WKL, ATH, SM, NF, ZH, GW, DS, LA, DL; Data Curation, VN, KKi, HBZ, MEB, RH, PK, ZS, SBS, AKS, LSK, JO-M, BB, JZ, WKL, SM, NF, ZH, GW, DS, LA, DL; Writing – Original Draft, VN, OS, JO-M; Writing – Review & Editing, all authors except KKo, BB, JZ, WKL, SM, NF, GW, DS, LA, and DL; Visualization, VN, KKi, MEB, RH, IP; Supervision, SBS, JD, OS, AKS, LSK, JO-M; Project Administration, SBS, JD, OS, AKS, LSK, JO-M, ATH, GW, DS, LA, DL; Funding Acquisition, OS, AKS, LSK, JO-M.

## Competing Interest Statement

VN, KKi, HBZ, AKS, LSK, and JO-M are co-inventors on a provisional patent application relating to methods of stratifying and treating IBD. SBS reports scientific advisory board participation for Pfizer, Lilly, IFM Therapeutics, Merck, Pandion, and Takeda; grant support from Pfizer, Novartis, Amgen, and Takeda; and consulting for Hoffmann-La Roche and Amgen. AKS reports compensation for consulting and/or scientific advisory board membership from Honeycomb Biotechnologies, Cellarity, Repertoire Immune Medicines, Ochre Bio, Parabilis Medicines, Passkey Therapeutics, Relation Therapeutics, Danaher, Conquest Technologies, IntrECate Biotherapeutics, and Dahlia Biosciences, unrelated to this work. AKS has also received research support from the Gates Foundation, the Moore Foundation, the Pew-Stewart Trust, Fondation MIT, the Chan Zuckerberg Initiative, Novo Nordisk, IBM, Wellcome Leap, Break Through Cancer, Coca-Cola, UK Research and Innovation, St. Jude Children’s Research Hospital, and the NIH, unrelated to this work. LSK serves on the scientific advisory boards of HiFiBio and Mammoth Biosciences. LSK reports research funding from Kymab Limited, Magenta Therapeutics, Bluebird Bio, and Regeneron Pharmaceuticals; consulting fees from Equillium, Forty Seven Inc., Novartis, EMD Serono, Gilead Sciences, Vertex Pharmaceuticals, and Takeda Pharmaceuticals; and grants and personal fees from Bristol Myers Squibb managed under an agreement with Harvard Medical School. JO-M reports compensation for consulting services from Tessel Biosciences, Radera Biotherapeutics, and Passkey Therapeutics. WKL, ATH, SM, ZH and NF are employees and stockholders of Regeneron Pharmaceuticals. GW reports research grants from Bristol Myers Squibb, Takeda, Pfizer, and GE, and service on a Data and Safety Monitoring Board for AbbVie. DS reports research support from Nestlé Health and Abbott. All other authors declare that they have no competing interests.

## Data Availability

The cell-by-gene matrices will be available for download with the peer-reviewed version of this manuscript and are currently viewable in the Broad Single Cell Portal (SCP) under study accession numbers
644 SCP3737 (blood), SCP3739 (total CD3 and NK cells from gut), and SCP3741 (CD4 T cells from gut).

## Acknowledgements

We thank the patients and their families for participating in and contributing to this study. We thank Kathy McConville and Sarah Mbonde for identifying and recruiting patients for PREDICT. VN was supported by the International Mobility of Research, Technical and Administrative Staff of Research Organizations program (CZ.02.2.69/0.0/0.0/18_053/0016981). Computational resources were provided by the e-INFRA CZ project (ID:90254), supported by the Ministry of Education, Youth and Sports of the Czech Republic. IP, JV, KKo, and JD are grateful for support from the Czech Science Foundation (26-21955S), the Foundation ACTERIA–ACTERIA Early Career Research Prize awarded by EFIS, the Ministry of Education, Youth and Sports of the Czech Republic grant ERC CZ (LL23/15), and the Programme Johannes Amos Comenius under the Ministry of Education, Youth and Sports of the Czech Republic (project no. CZ.02.01.01/00/22_008/0004597). LN was supported by the National Institute of General Medical Sciences (T32GM007753 and T32GM144273). SBS was supported by NIH grants P30 DK034854 and RC2 DK122532 and by the Helmsley Charitable Trust. OS was supported by Czech Health Research Council grant NW26-05-00027. AKS was supported, in part, by the Searle Scholars Program, the Beckman Young Investigator Program, a Sloan Fellowship in Chemistry, and the NIH (5U24AI118672 and 2R01HL095791). LSK was supported by NIH grants P01HL158504, R01HL095791, and U19AI051731, and by the Helmsley Charitable Trust. JO-M is a New York Stem Cell Foundation–Robertson Investigator. JO-M was supported by the AbbVie–Harvard Medical School Alliance; the AGA Research Foundation’s AGA–Takeda Pharmaceuticals Research Scholar Award in IBD (AGA2020-13-01); the Leona M. and Harry B. Helmsley Charitable Trust; The Pew Charitable Trusts Biomedical Scholars Program; the Broad Next Generation Award; the Chan Zuckerberg Initiative Pediatric Networks; The Mathers Foundation; The Kenneth Rainin Foundation; the Massachusetts Consortium for Pathogen Readiness; The New York Stem Cell Foundation; NIH grants R01HL162642, R01DE031928, and P30DK034854; the NIH CIDER Network; Project NextGen; and The Cell Discovery Network, a collaboration funded by The Manton Foundation and The Warren Alpert Foundation at Boston Children’s Hospital. The content is solely the responsibility of the authors and does not necessarily represent the official views of the National Institute of General Medical Sciences or the National Institutes of Health.

---

**Supplementary Figure 1.**
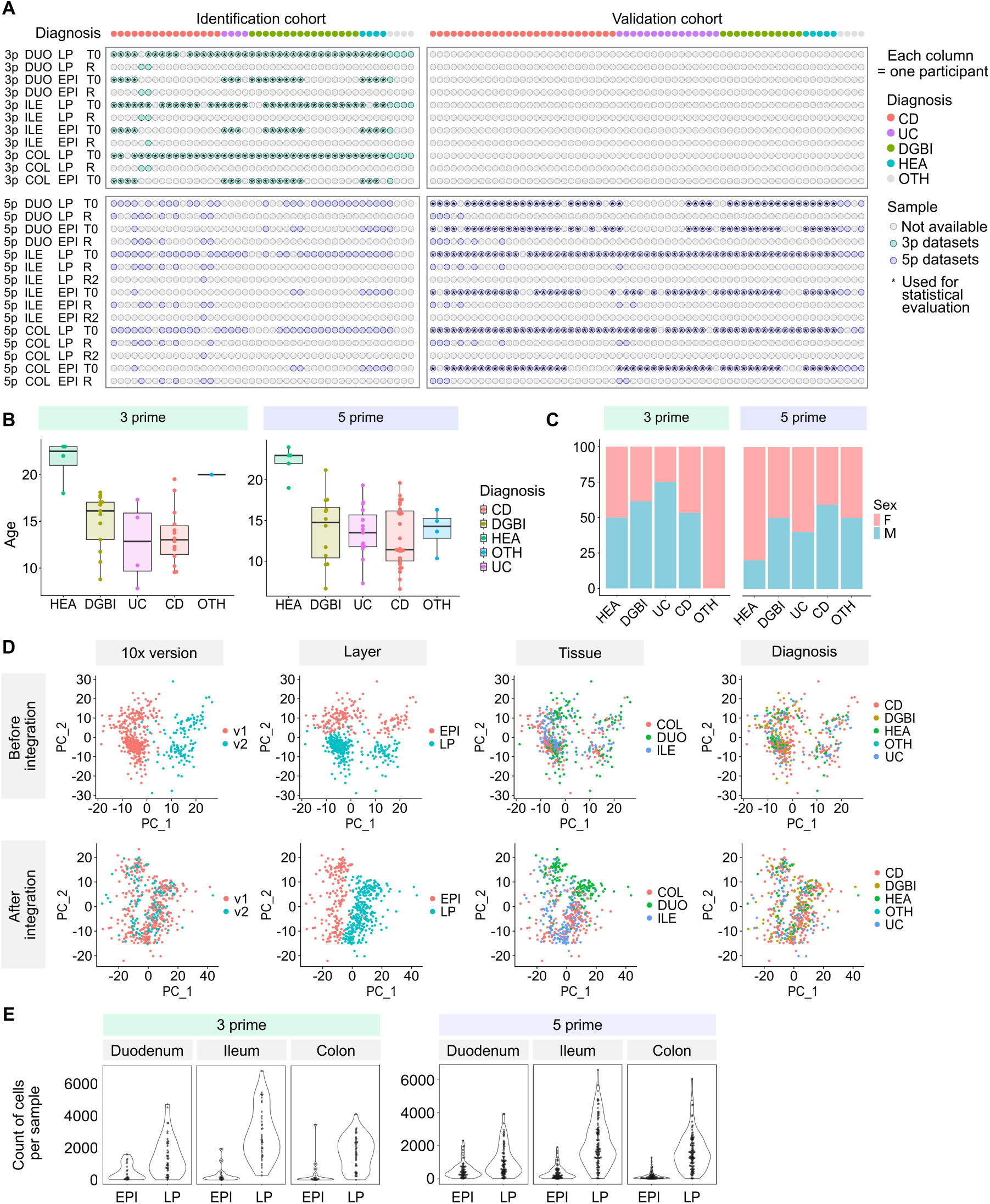
Overview of the PREDICT study cohort and gut atlas datasets. **(A)** Schematic representation of participants in the PREDICT study and samples included in the gut atlas. Each column represents one participant, and rows indicate which samples from the same participant were included in the analysis. DUO – duodenum; ILE – ileum; COL – colon; LP – lamina propria; EPI – epithelium; T0 – time point at diagnosis; R – repeated visit. **(B)** Boxplots showing the age distribution of participants in the 3p and 5p datasets. Boxplots show the median (center line), interquartile range (box), and whiskers extending to 1.5× the interquartile range. CD – Crohn’s disease; DGBI – disorders of gut–brain interaction; UC – ulcerative colitis; HEA – healthy; OTH – other. 3p: n = 36, 5p: n = 63. **(C)** Bar plots showing the sex distribution of participants in the 3p and 5p datasets. Donor counts are as in (B). **(D)** Sample-level PCA of 5p T cell samples before (top panels) and after (bottom panels) integration over batch and donor using STACAS. Each dot represents one sample. Samples are colored by 10x chemistry version, tissue layer, tissue location, and diagnosis. **(E)** Cell counts per sample for samples derived from the epithelial layer (EPI) or lamina propria (LP).

**Supplementary Figure 2.**
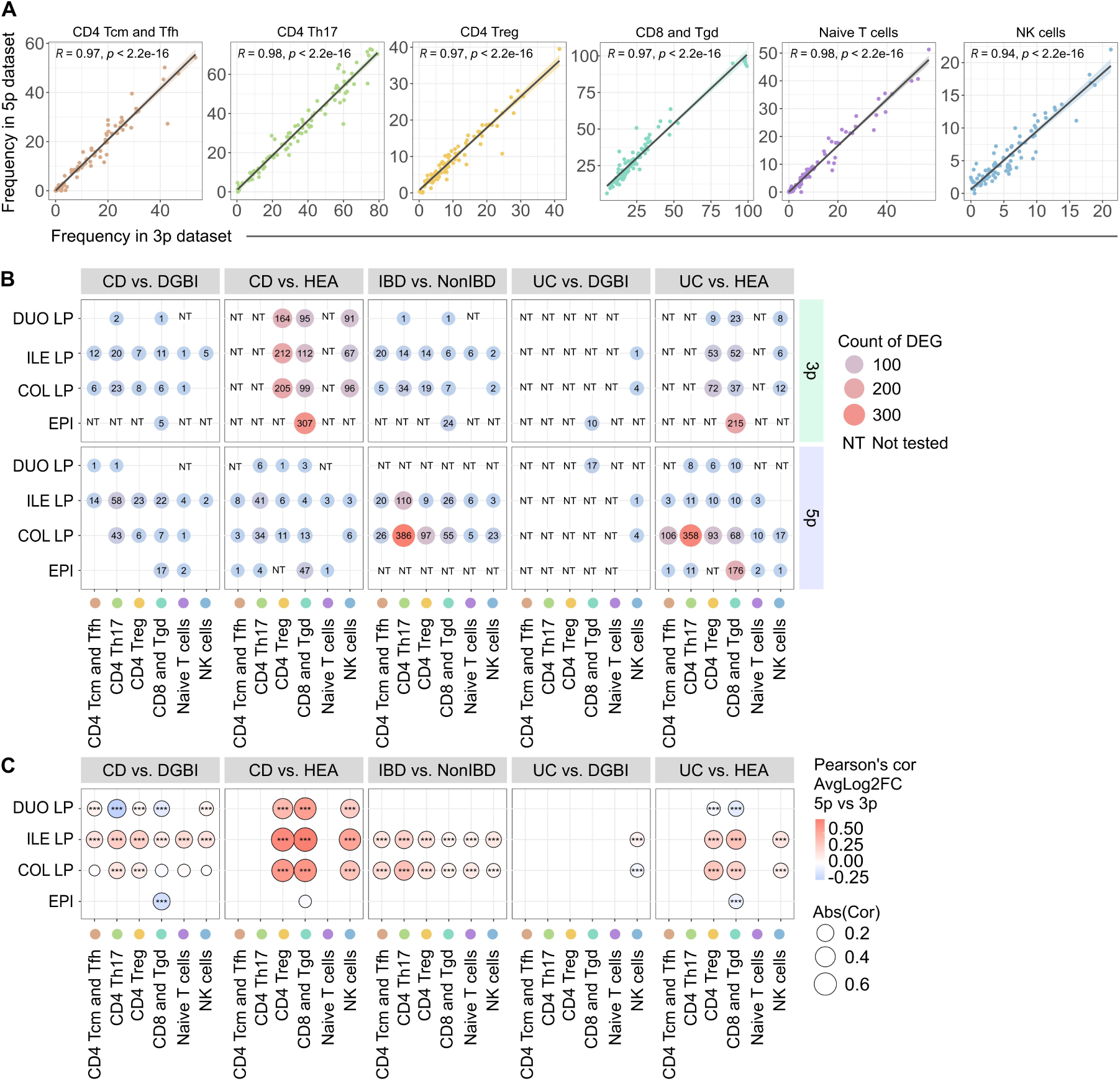
Comparison of 3p and 5p PREDICT gut atlas datasets. **(A)** Correlation of the frequencies of cell subsets from Fig. 1D in the PREDICT 3p dataset and the PREDICT 5p dataset. Included are samples from the same tissue and the same participant analyzed independently using the two methods. The solid line represents the linear regression fit, and the shaded area indicates the 95% confidence interval. **(B)** Dot plot showing the number of differentially expressed genes passing the threshold of adjusted p value (p.adj) < 0.05 in the indicated cell populations, tissues, and contrasts. The size and color of the dots represent gene counts. NT – not tested in cases where fewer than three pseudobulk samples were available for any condition. **(C)** Dot plot showing the correlation between average log₂ fold changes (avgLog₂FC) for all tested genes in the indicated population, tissue, and condition in the 3p versus 5p datasets. The color and size of the dots represent the correlation coefficient, and stars indicate statistical significance. P values were calculated using two-tailed Pearson correlation tests and adjusted for multiple comparisons using Bonferroni correction.

**Supplementary Figure 3.**
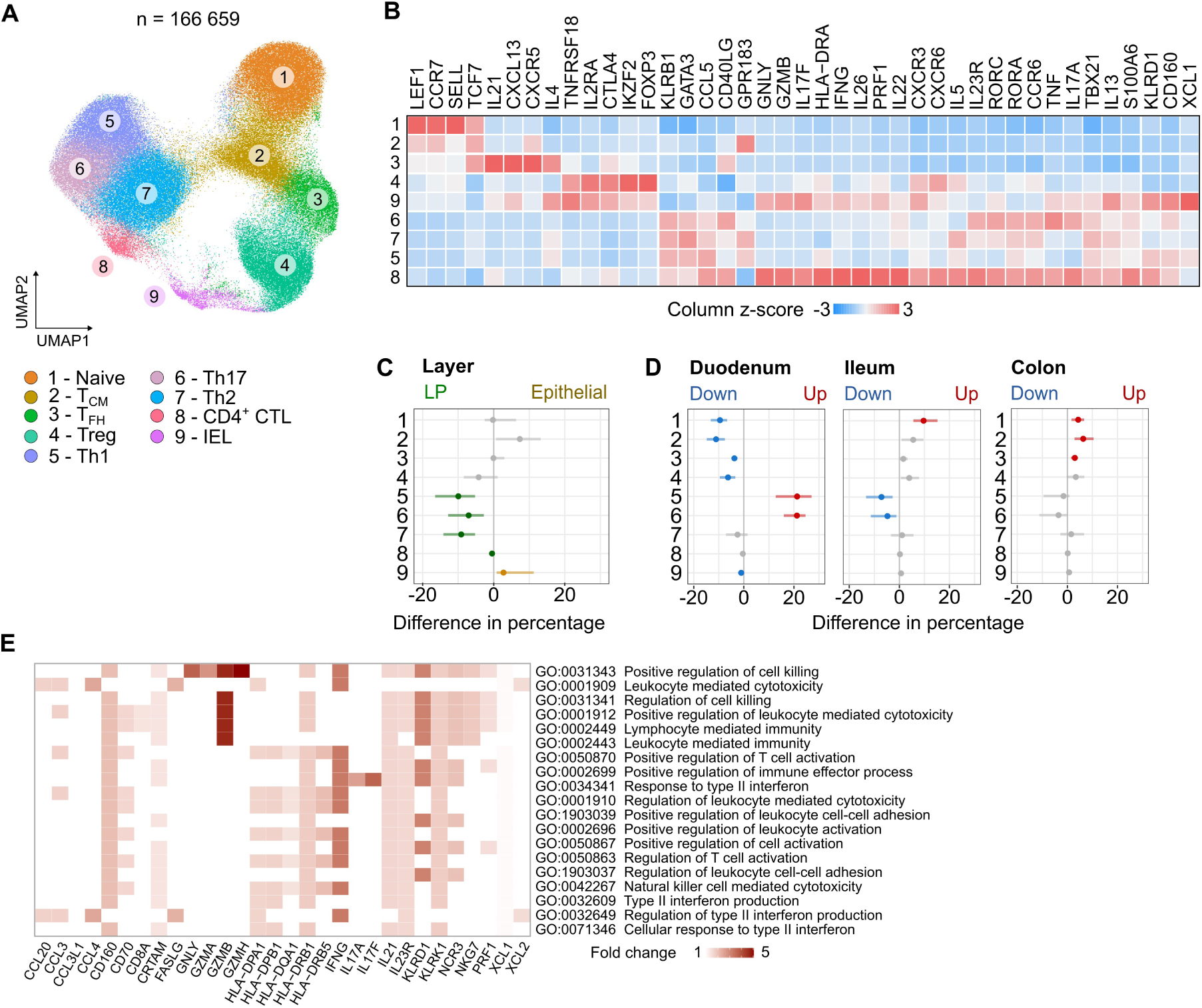
Characterization of gut CD4⁺ T cell subsets in the PREDICT 3p dataset. **(A)** UMAP projection of CD4⁺ T lymphocytes from the PREDICT 3p gut dataset (n = 166,659 cells from 39 donors). Cells are colored by manual cluster annotations. **(B)** Heatmap showing selected marker genes defining the clusters shown in (A). Colors represent the row-scaled z-score of average gene expression per cluster. **(C–D)** Forest plots showing changes in the proportions of clusters shown in (A) in samples derived from different gut layers, lamina propria (LP) versus epithelium (C), and from different gut locations (D). **(E)** Gene set enrichment analysis (GSEA) of differentially expressed genes (DEGs) in CD4 CTL cells compared to other gut CD4⁺ T cells. DEGs were identified using the *FindMarkers* function, and the top 50 genes with avg_log₂FC > 1.5 were used for GSEA with *enrichGO* from *clusterProfiler*. The top 20 pathways are shown; color represents the average log₂ fold change in CD4 CTL cells relative to other CD4⁺ T cells.

**Supplementary Figure 4.**
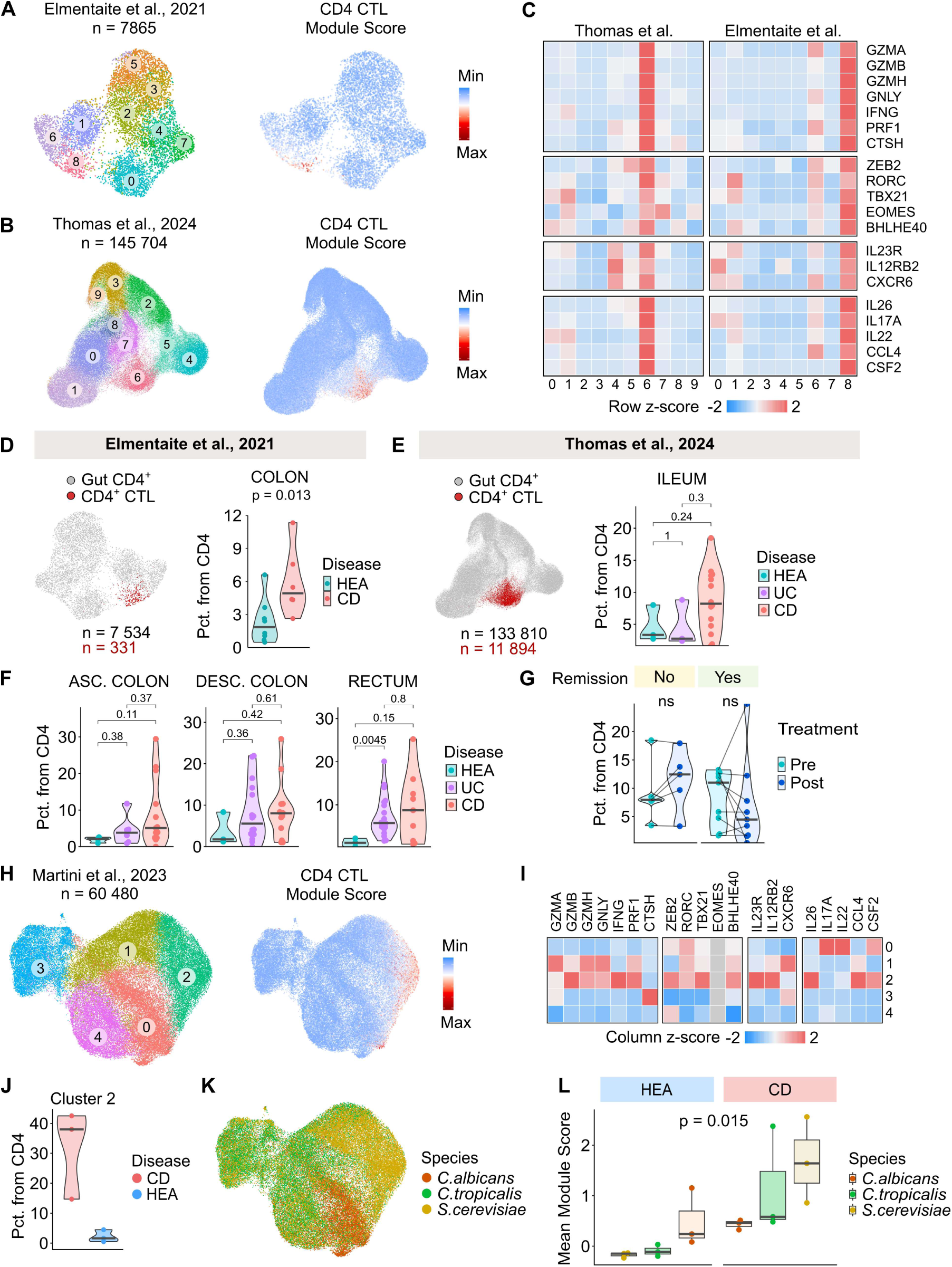
Validation of the CD4 CTL transcriptional program in independent human datasets. (A–B) UMAP projections of CD4⁺ T cells from the Elmentaite et al. dataset **(A)** and the Thomas et al. dataset **(B)**. Left panels show CD4⁺ T cell clusters, and right panels show the module score calculated from the 187 top markers of the CD4 CTL cluster from the 5p PREDICT dataset. (A) n = 7,865 cells from 14 donors; (B) n = 145,704 cells from 41 donors. **(C)** Heatmap showing selected marker genes defining the clusters shown in (A) and (B). Colors represent the row-scaled z-score of average gene expression per cluster. **(D)** Proportions of CD4 CTL cells in participants from the Elmentaite *et al.* cohort. *Left*: UMAP projection of CD4⁺ T cells. *Right*: Violin plots showing the percentage of CD4 CTL cells among total CD4⁺ T cells. P values were calculated using a two-tailed Mann–Whitney U test. Horizontal bars indicate medians. **(E)** Proportions of CD4 CTL cells in ileum of participants from the Thomas *et al.* cohort. *Left*: UMAP projection of CD4⁺ T cells. *Right*: Violin plots showing the percentage of CD4 CTL cells among total CD4⁺ T cells. P values were calculated as in (D). Bars indicate medians. **(F)** Same as E but for ascending colon (left), descending colon (middle) and rectum (right). **(G)** Percentage of CD4 CTL cells among total CD4⁺ T cells in ileal samples of Thomas et. al cohort stratified by remission status and treatment. Lines connect the same donors before and after treatment. P values were calculated using paired Wilcoxon signed-rank test. **(H)** UMAP projection of CD4⁺ T cells from the Rios-Martini et al. dataset. Left panel shows clustering, right panel shows the module score calculated from the 187 top markers of the CD4 CTL cluster from the 5p PREDICT dataset. n = 60,480 cells from 6 donors. **(I)** Heatmap showing selected marker genes defining the clusters shown in (H). Colors represent the column-scaled z-score of average gene expression per cluster. **(J)** Quantification of the proportions of cluster 2 within all clusters in donors with CD and healthy donors. **(K)** The same UMAP as in (H) colored by the origin of yeast antigen. Cells were stimulated ex vivo with lysates from the indicated yeasts. **(L)** Box plots showing the mean module score calculated from the 187 top markers of the CD4 CTL cluster from the 5p PREDICT dataset in the Rios-Martini et al. dataset. Boxplots show the median (center line), interquartile range (box), and whiskers extending to 1.5× the interquartile range.

**Supplementary Figure 5.**
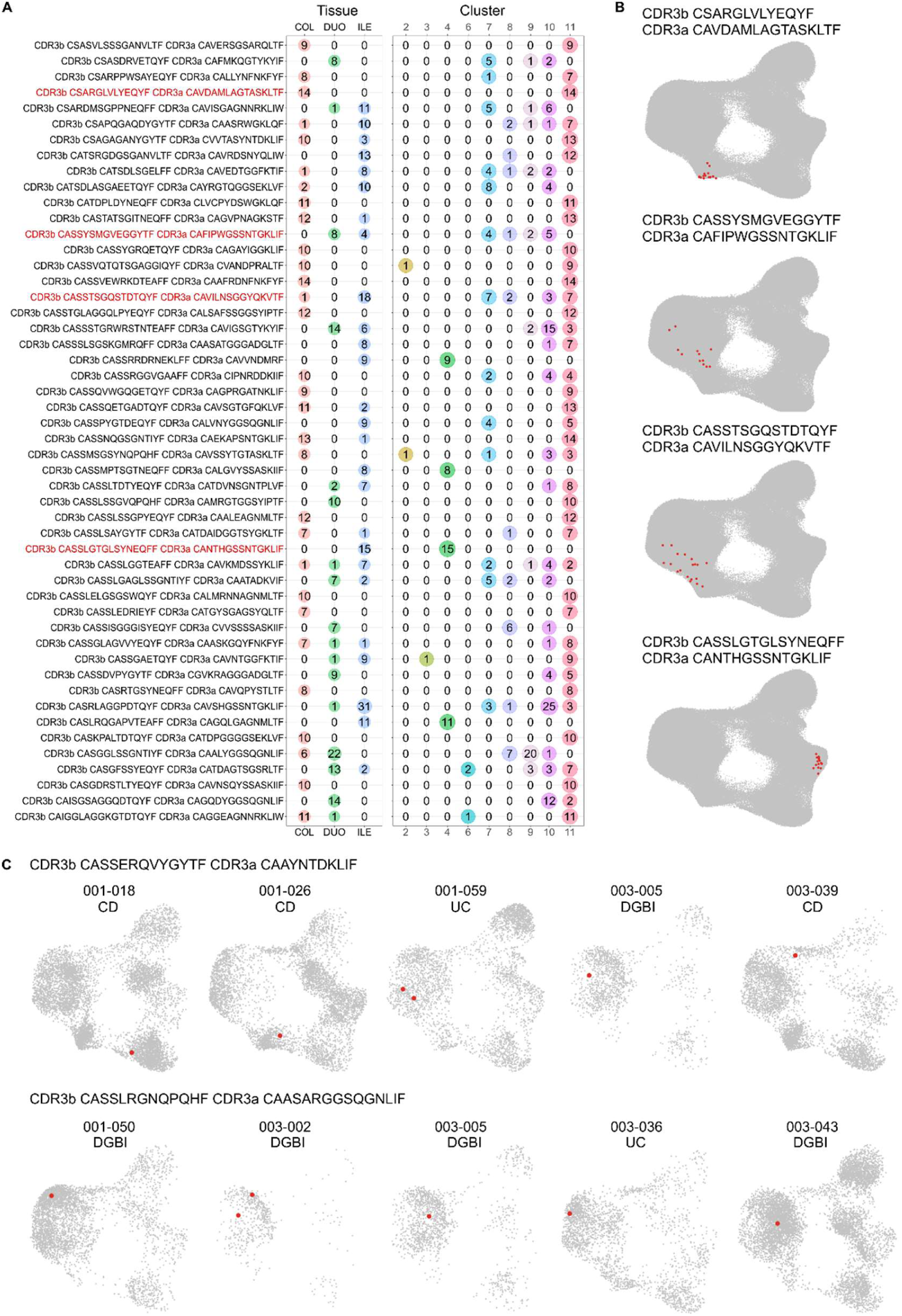
Clonal expansion and distribution of CD4⁺ T cell clones in the gut. **(A)** List of the top 50 most expanded T cell clones, their CDR3 sequences, degree of expansion across different gut locations, and phenotypic assignment within the clusters shown in Fig. 2A. TCRs highlighted in red are visualized in (B). **(B)** UMAP projection as in Fig. 2A showing selected expanded T cell clones exhibiting distinct phenotypes. **(C)** The two most shared TCR clones with identical TCR receptor sequences detected across the highest number of participants.

**Supplementary Figure 6.**
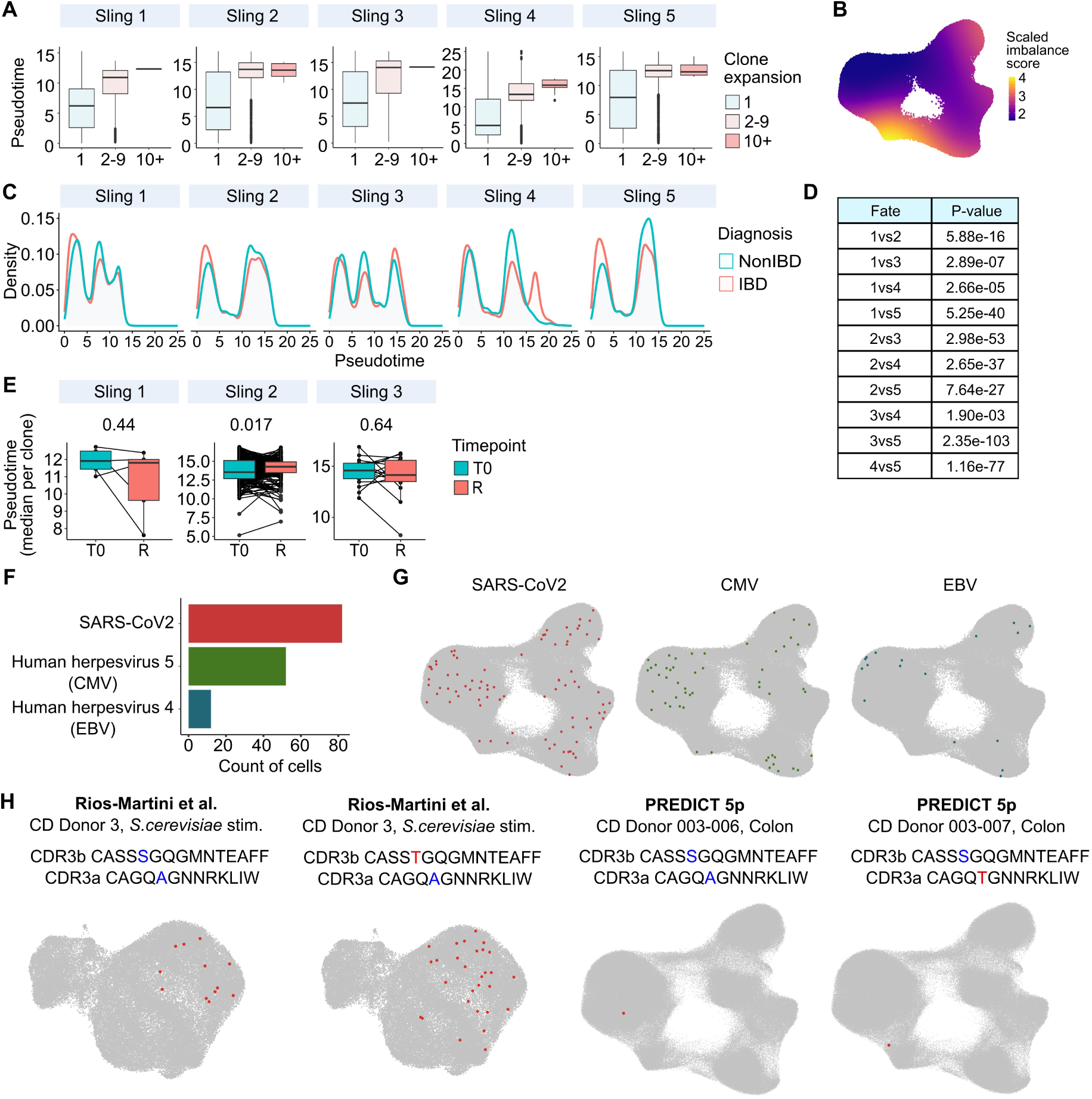
Clonal dynamics, pseudotime progression, and antigen specificity of gut CD4⁺ T cells. **(A)** Boxplots showing the average pseudotime per clone grouped by clone expansion. Boxplots show the median (center line), interquartile range (box), and whiskers extending to 1.5× the interquartile range. **(B)** UMAP projection as in Fig. 2A, colored by scaled imbalance score estimated using the condiments package. Higher scores indicate greater differences in cell density between healthy donors and donors with Crohn’s disease (CD), ulcerative colitis (UC), and DGBI. **(C)** Density of CD4⁺ T cells from IBD (CD and UC) and non-IBD (healthy, DGBI) donors along inferred developmental trajectories. **(D)** Table of pairwise P values from condiments differential fate selection analysis (fateSelectionTest) comparing lineage choice probabilities between IBD and non-IBD CD4⁺ T cells across inferred developmental trajectories. **(E)** Boxplots showing the median pseudotime per clone grouped by time point of sampling. T0 – at diagnosis; R – 3 months after diagnosis. Boxplots show the median (center line), interquartile range (box), and whiskers extending to 1.5× the interquartile range. P value was calculated using a two-tailed paired Wilcoxon signed-rank test. **(F-G)** Similarity of CDR3 sequences of the TCRβ chain from cells in the PREDICT 5p dataset was compared with known CDR3 TCRβ sequences binding epitopes curated in the Immune Epitope Database. Only matches with a similarity score > 0.99 were considered. **(F)** Bar plots showing the number of cells with matching CDR3 TCRβ sequences corresponding to SARS-CoV-2, human herpesvirus 5 (CMV, cytomegalovirus), or human herpesvirus 4 (EBV, Epstein–Barr virus). **(G)** UMAP projection as in Fig. 2A highlighting cells with CDR3 TCRβ sequences matching those of SARS-CoV-2, CMV, or EBV. **(H)** Projection of a clonotype shared in PREDICT and Rios Martini cohorts.

**Supplementary Figure 7.**
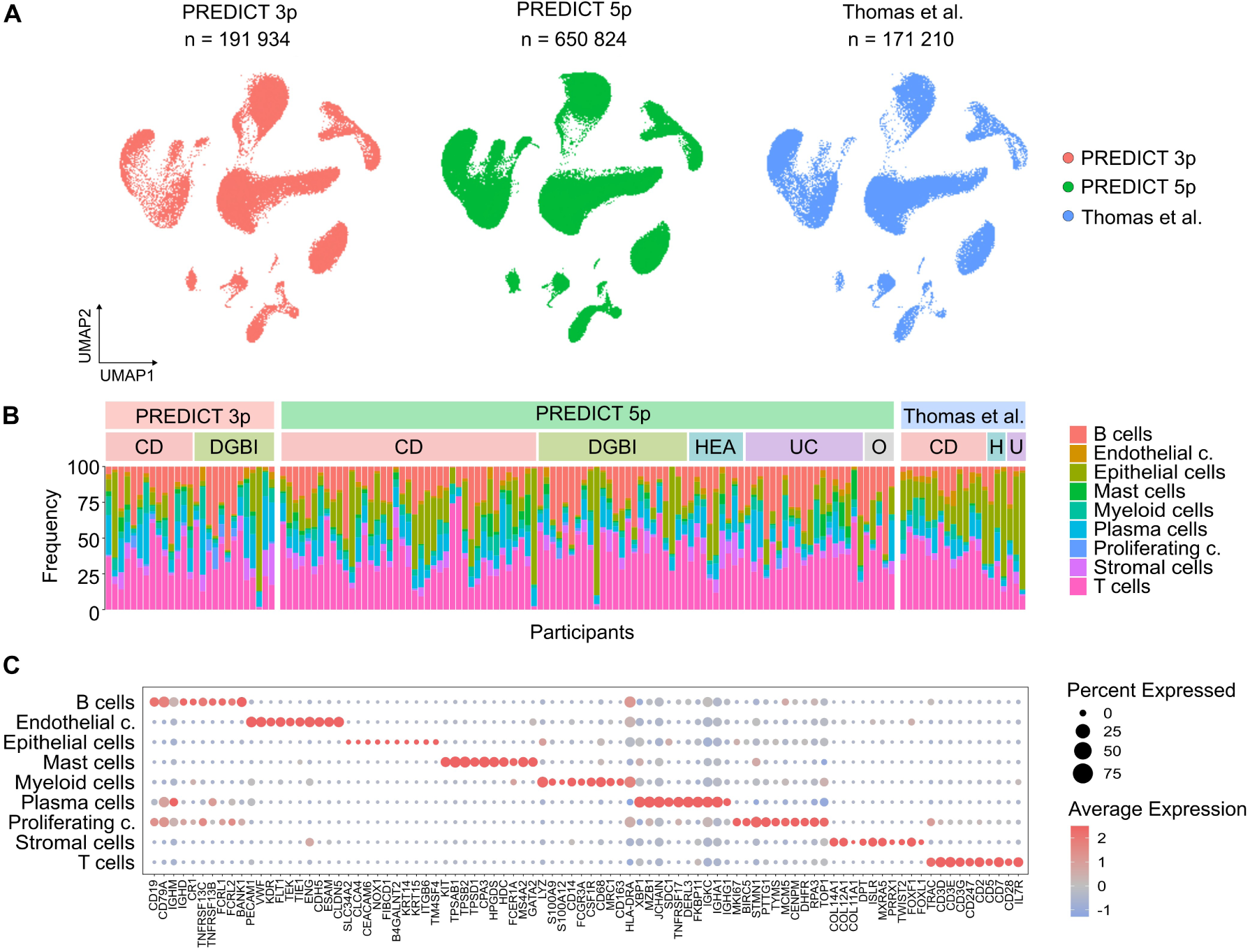
Cohort-specific representation of major immune cell populations. **(A)** UMAP projection as in Fig. 4A, split by dataset of origin. **(B)** Bar plot showing the proportions of the main cell populations defined in Fig. 4A across participants from the three cohorts. CD - Crohn’s disease, UC - ulcerative colitis, DGBI - disorders of gut-brain interaction, HEA - healthy. **(C)** Dot plot showing the main marker genes defining the populations presented in Fig. 4A.

**Supplementary Figure 8.**
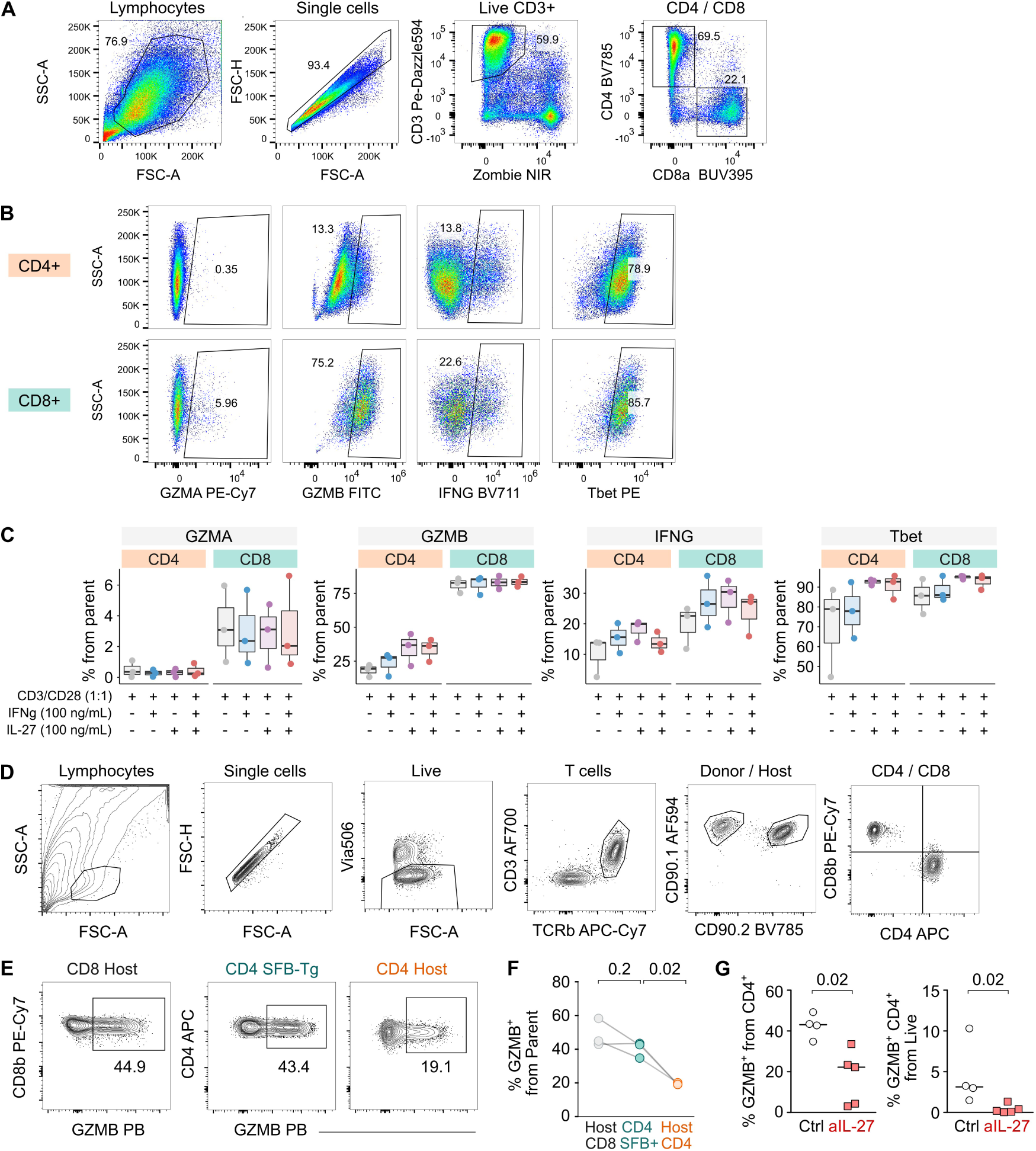
IL-27 is required for acquisition of cytotoxic phenotype in CD4^+^ T cells. (A-C) PBMCs from 3 independent healthy donors were stimulated in vitro with anti-CD3/CD28 beads for 70 hours. Recombinant human IFN-γ and/or IL-27 were added at a final concentration 100 ng/mL. Expression of GZMA, GZMB, IFNG and Tbet in CD4^+^ T cells and CD8^+^ T cells post stimulation were measured using flow cytometry. **(A)** Representative gating strategy for identification of CD4^+^ and CD8^+^ T cells. **(B)** Representative gating strategy for identification of GZMA, GZMB, IGNF and Tbet positive T cells. **(C)** Quantification of the percentage of CD4^+^ and CD8^+^ T cells positive for GZMA, GZMB, IGNF and Tbet under different stimulation conditions. Boxplots show the median (center line), interquartile range (box), and whiskers extending to 1.5× the interquartile range. n = 3 biological replicates. **(D)** Representative gating strategy for analysis of intestinal T cells from the adoptive transfer mouse model. **(E)** Representative flow cytometry plots showing gating of GZMB⁺ T cells among host CD8⁺ T cells, donor SFB-Tg CD4⁺ T cells, and host CD4⁺ T cells. **(F)** Quantification of GZMB⁺ T cells among host CD4⁺ T cells, donor SFB-Tg CD4⁺ T cells, and host CD8⁺ T cells. P-values were calculated using two-tailed paired t-test. **(G)** Quantification of the percentage of GZMB⁺ CD4⁺ T cells gated from total CD4⁺ T cells (left), or from live cells (right). Bars indicate medians. SFB – segmented filamentous bacteria. n = 9 mice in two experiments.

**Supplementary Figure 9.**
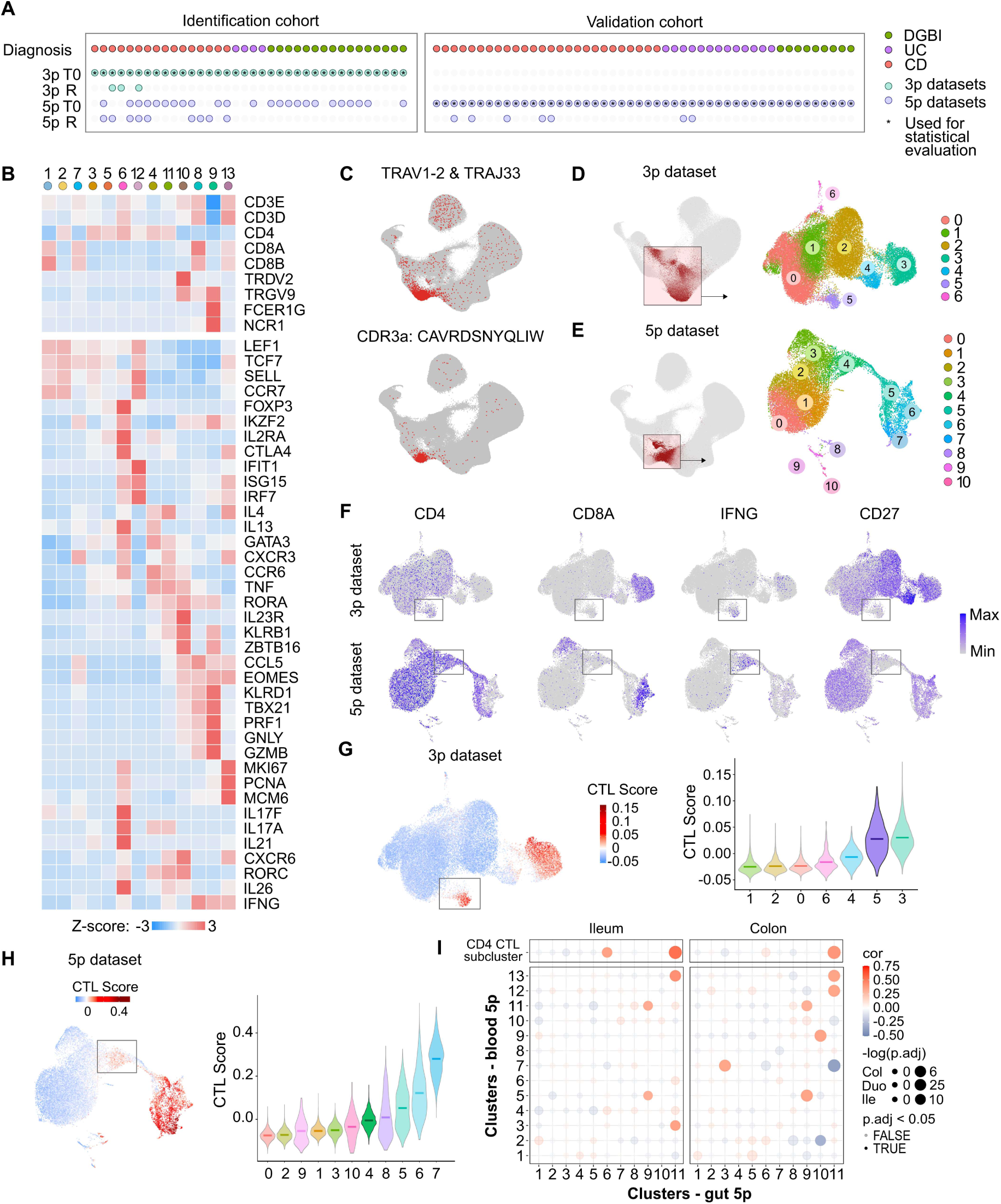
Composition and reclustering of the peripheral blood T cell atlas in the PREDICT study. **(A)** Schematic representation of participants in the PREDICT study and samples included in the blood atlas. Each column represents one participant, and rows indicate which samples from the same participant were included in the analysis. T0 – time point at diagnosis; R – repeated visit; CD – Crohn’s disease; DGBI – disorders of gut–brain interaction; UC – ulcerative colitis. **(B)** Heatmap showing selected marker genes defining clusters from the PREDICT 3p dataset shown in Fig. 5A. Colors represent the row-scaled z-score of average gene expression per cluster. The CD4 CTL column represents CD4 CTL cells derived from reclustering. **(C)** The same UMAP projection as in Fig. 5B showing the TCR rearrangements specific for MAIT cells. **(D-E)** Reclustering of Th17-like and Treg-like clusters from the peripheral blood T cell atlas of the 5p dataset shown in Fig. 5B (D) and from the 3p dataset shown in Fig. 5A (E). Clusters selected for reclustering (left panels) were subjected to new normalization, scaling, integration, and clustering, resulting in refined clusters (right panels). **(F)** Feature plots showing expression of selected markers in UMAP projections after reclustering. **(G-H)** Visualization (left) and quantification (right) of the module score calculated from the 187 top markers of the CD4 CTL cluster from the 5p PREDICT dataset after reclustering. **(I)** Dot plot showing the correlation between frequencies of clusters from the gut 5p dataset shown in Fig. 2A and frequencies of clusters from the blood 5p dataset shown in Fig. 5B. Dot color represents the correlation coefficient and dot size represents –log(adjusted p value). P values were calculated using two-tailed Pearson correlation tests and adjusted for multiple comparisons using Bonferroni correction.

**Supplementary Figure 10.**
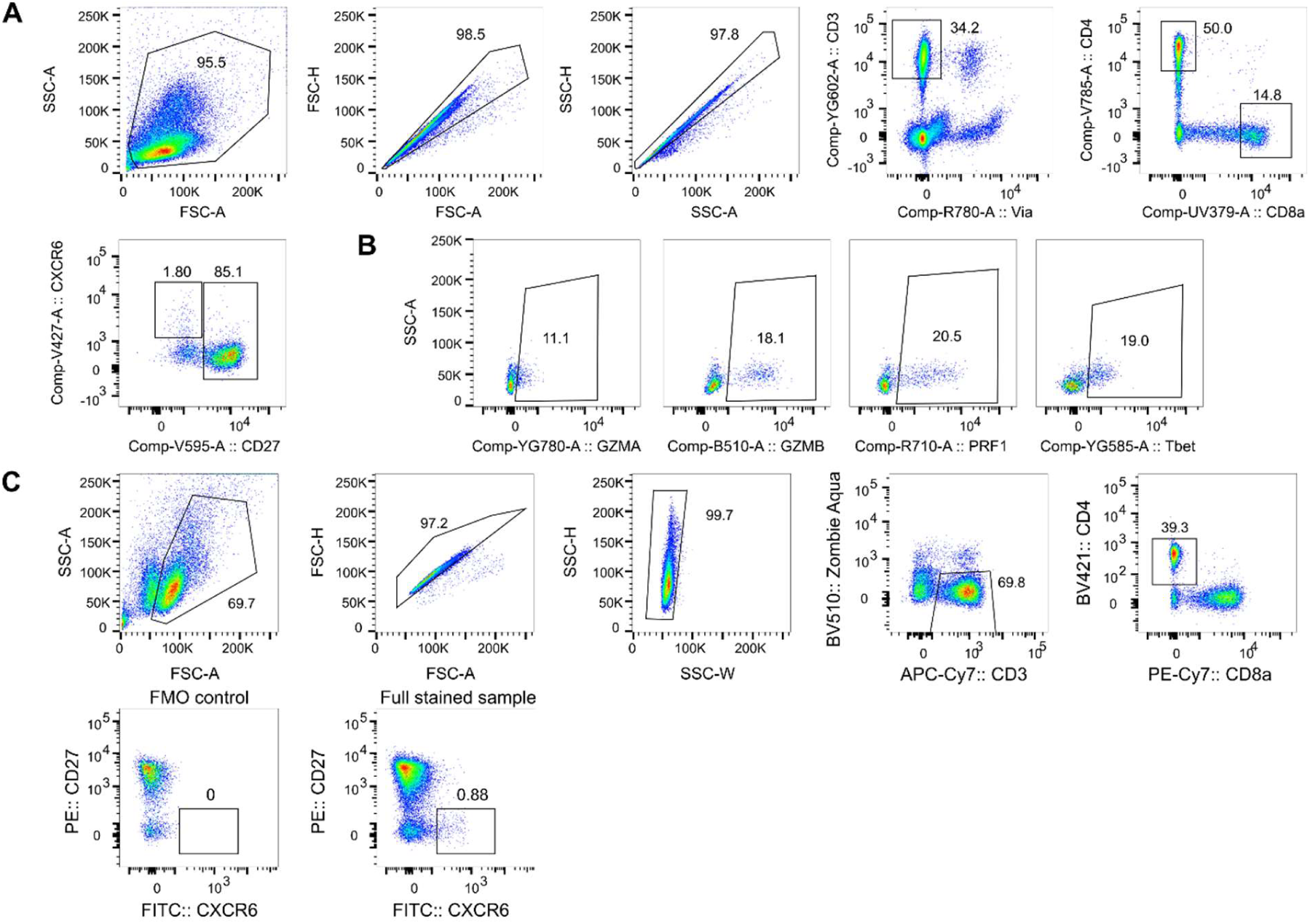
Flow cytometry gating strategies for identification, functional profiling, and sorting of CD4 CTL cells. **(A)** Representative gating strategy for identification of CD4 CTL cells related to Fig. 6A. **(B)** Representative gating strategy for identification of cells positive for GZMA, GZMB, PRF1, and T-bet related to Fig. 6C. **(C)** Representative gating strategy for sorting CD4 CTL cells for the scRNA-seq validation experiment related to Fig. 6F.

**Supplementary Table S1.**
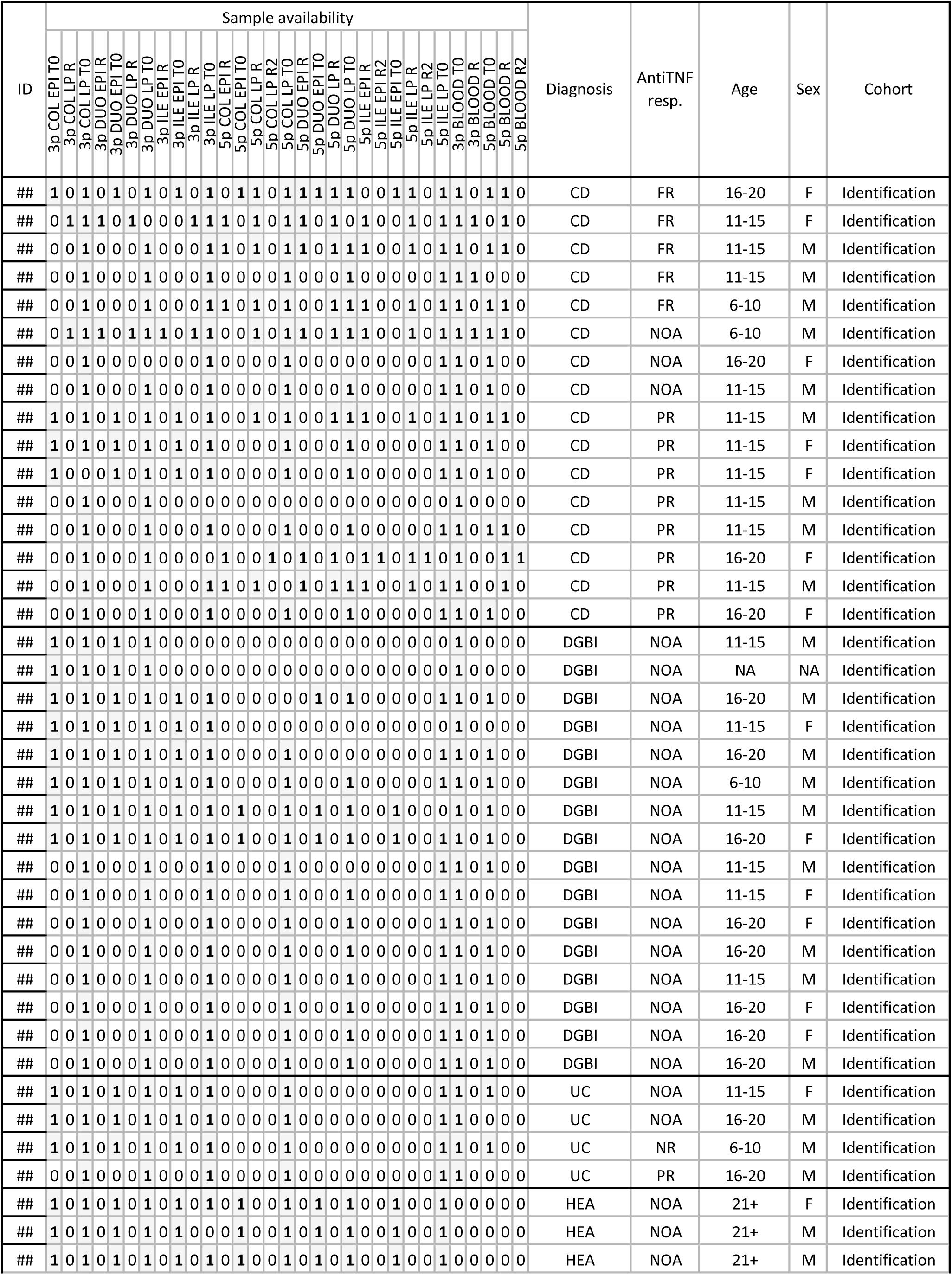
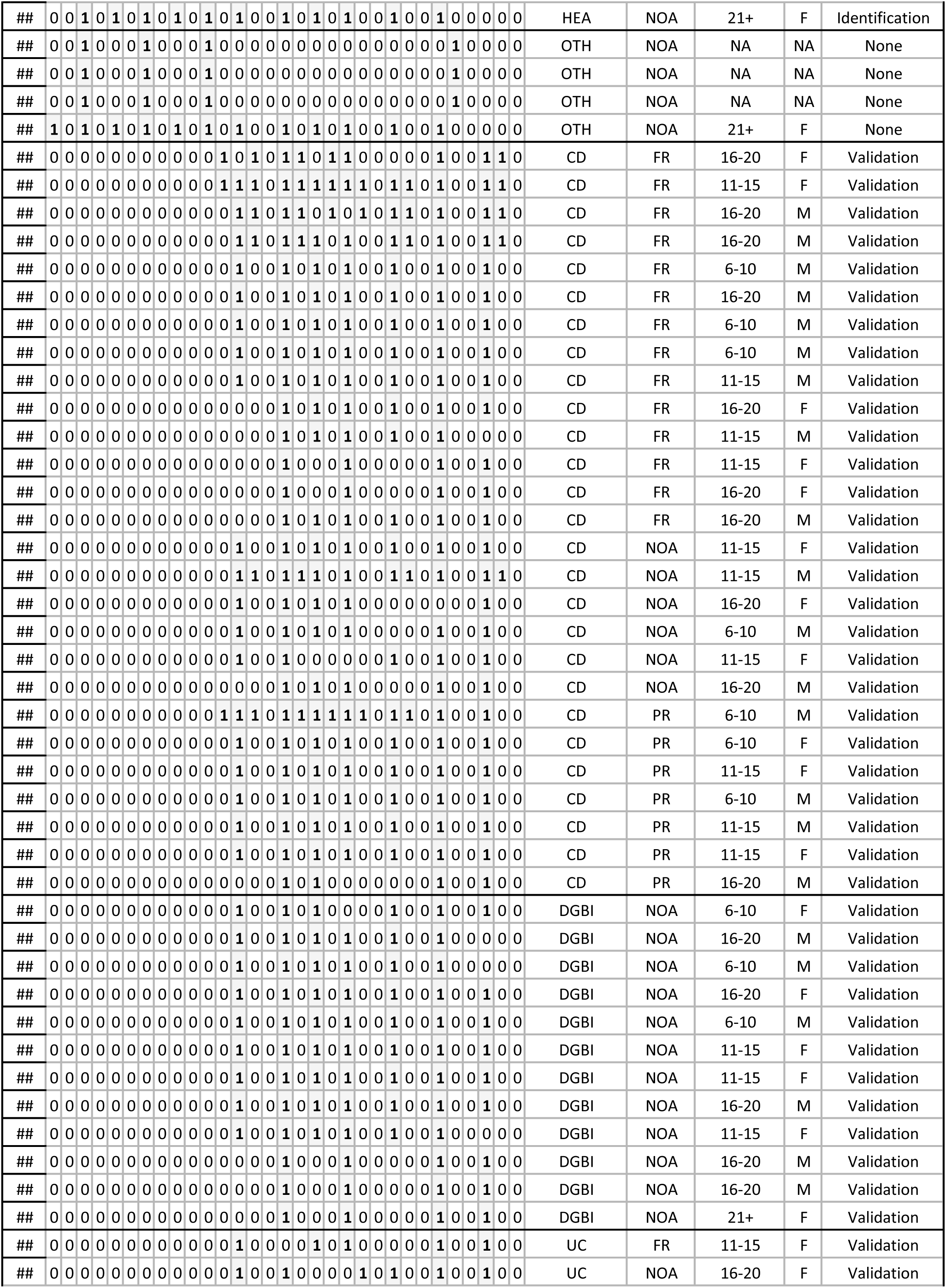
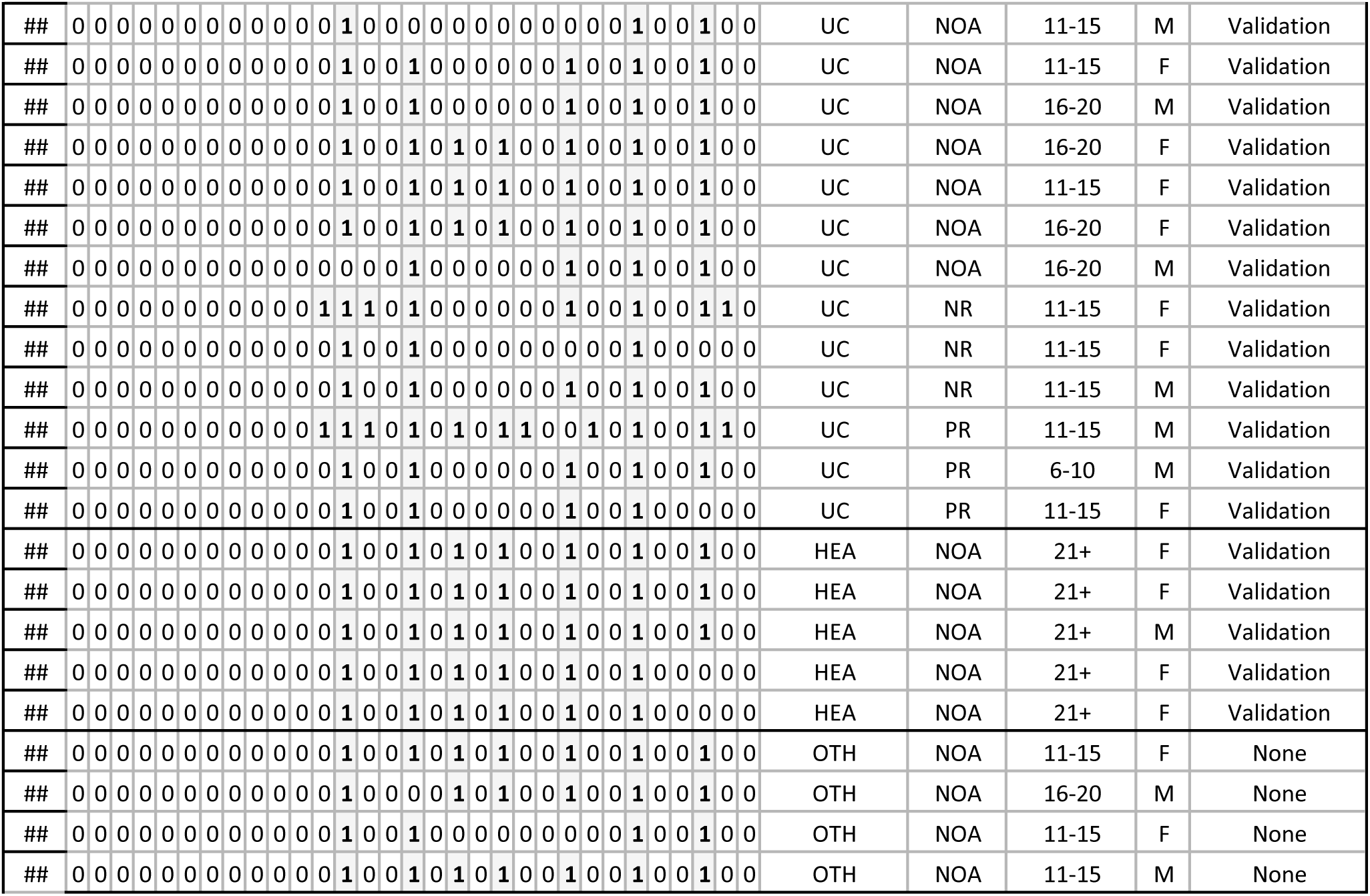
List of participants of PREDICT cohort and samples used in this study.

**Supplementary Table S2.**
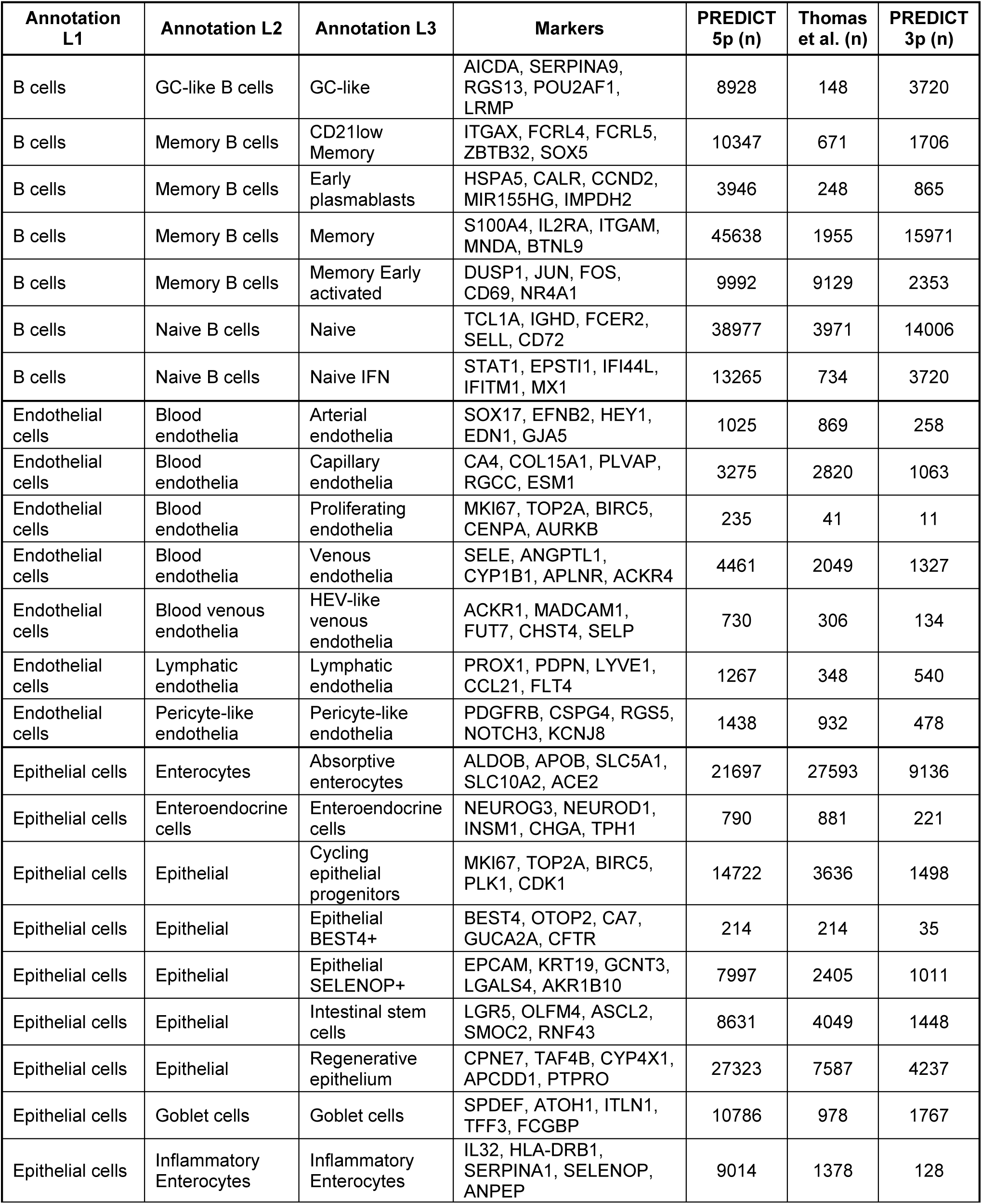
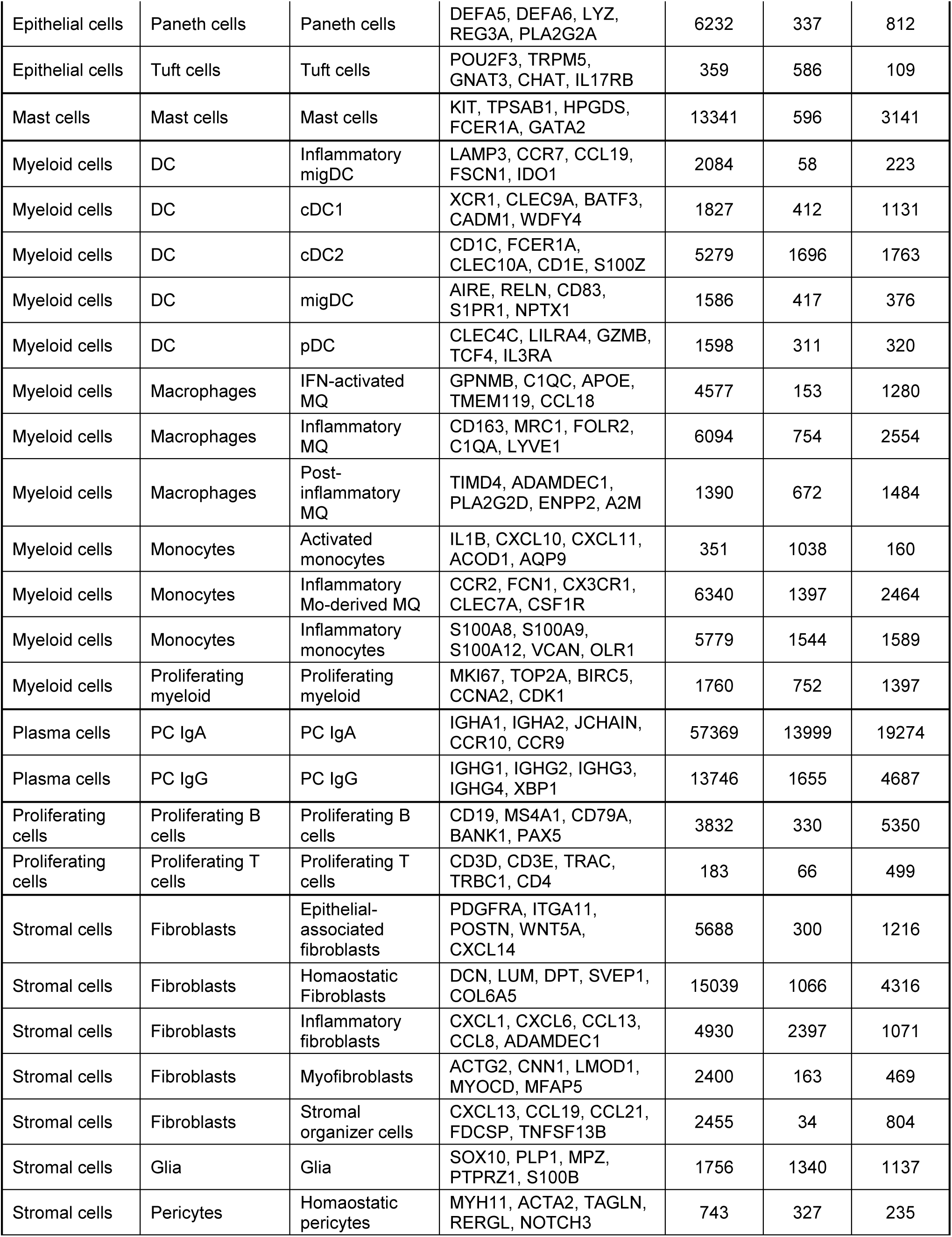
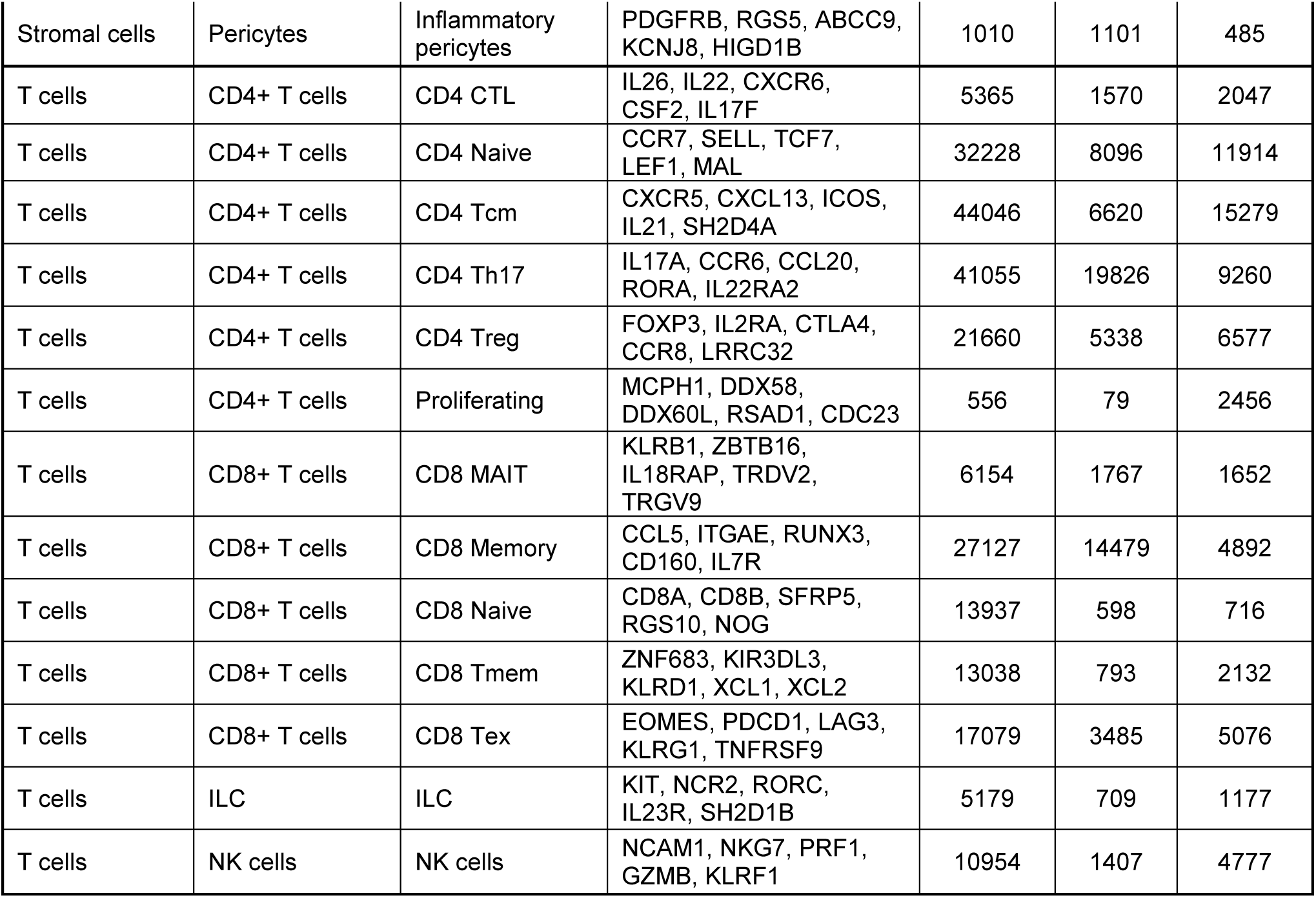
List of annotation of all cell types and cell subsets related to Fig. 4.

**Supplementary Table S3.** List of participants whose PBMC samples were used for validation experiment with scRNA-seq profiling of sorted CD4 CTL.

| Age | Sex | Severe CD Criteria |
| --- | --- | --- |
| 11-15 | F | perianal, fistulizing, stricturing Crohn's, developed antibodies to infliximab and now on humira |
| 11-15 | M | fistulizing Crohn's with perianal abscess |
| 16-20 | M | refractory to infliximab and stelara, now on skyrizi |
| 11-15 | M | refractory to infliximab and stelara, has JIA as an EIM |
| 16-20 | M | underwent ileocecal resection which revealed chronic, severely active ileocolitis with transmural inflammation and strictures |

**Supplementary Table S4.** Reagents used for flow cytometry and cell sorting of human cells.

| Name | Fluorochrome | Marker | Clone | Manufacturer | Cat | LOT | Dilution |
| --- | --- | --- | --- | --- | --- | --- | --- |
| **Panel 1** |  |  |  |  |  |  |  |
| BD Horizon™<br>BUV395 Mouse<br>Anti-Human CD8 | BUV395 | CD8a | RPA-T8 | BD | 563795 | 1097745 | 400x |
| BD Horizon™<br>BUV805 Mouse<br>Anti-Human CD14 | BUV805 | CD14 | M5E2 | BD | 612903 | 4250797 | 400x |
| Brilliant Violet<br>421™ anti-human<br>CD186 (CXCR6)<br>Antibody | BV421 | CXCR6 | K041E5 | BioLegend | 356013 | B342442 | 100x |
| Brilliant Violet<br>605™ anti-human<br>CD27 Antibody | BV605 | CD27 | O323 | BioLegend | 302829 | B391607 | 300x |
| BD Horizon™<br>BV711 Mouse<br>Anti-Human IFN-γ | BV711 | IFNγ | B27 | BD | 564039 | 1197770 | 300x |
| Brilliant Violet<br>785™ anti-human<br>CD4 Antibody | BV785 | CD4 | OKT4 | BioLegend | 317441 | B426990 | 400x |
| FITC anti-human/mouse<br>Granzyme B<br>Antibody | FITC | GZMB | GB11 | BioLegend | 515403 | B351810 | 300x |
| BD Pharmingen™<br>PerCP-Cy™5.5<br>Mouse Anti-Human IL-17A | PerCP-Cy5.5 | IL17A | N49-653 | BD | 560799 | 1131141 | 100x |
| PE anti-human T-bet Recombinant<br>Antibody | PE | Tbet | QA18A24 | BioLegend | 365903 | B412345 | 300x |
| PE/Dazzle™ 594<br>anti-human CD3<br>Antibody | Pe-Dazzle594 | CD3 | OKT3 | BioLegend | 317345 | B443448 | 400x |
| PE/Cyanine7 anti-human Granzyme<br>A Antibody | PE-Cy7 | GZMA | CB9 | BioLegend | 507221 | B360334 | 100x |
| Human IL-23R<br>APC-conjugated<br>Antibody | APC | IL23R | 218213 | R&D | FAB14001A | LZH0821071 | 100x |
| Alexa Fluor® 700<br>anti-human<br>Perforin Antibody | AF700 | PRF1 | B-D48 | BioLegend | 353323 | B332512 | 400x |
| Zombie NIR<br>Fixable Viability<br>Kit | NIR | Via |  | BioLegend | 423106 | B419021 | 1000x |
| **Panel 2** |  |  |  |  |  |  |  |
| BD Horizon™<br>BUV395 Mouse<br>Anti-Human CD8 | BUV395 | CD8a | RPA-T8 | BD | 563795 | 1097745 | 400x |
| BD Horizon™<br>BUV805 Mouse<br>Anti-Human CD14 | BUV805 | CD14 | M5E2 | BD | 612903 | 4250797 | 400x |
| Brilliant Violet 421™ anti-human Ki-67 Antibody | BV421 | Ki67 | Ki-67 | BioLegend | 350505 | B356739 | 400x |
| Brilliant Violet 605™ anti-human CD27 Antibody | BV605 | CD27 | O323 | BioLegend | 302829 | B391607 | 300x |
| BD Horizon™ BV711 Mouse Anti-Human IFN-γ | BV711 | IFNγ | B27 | BD | 564039 | 1197770 | 300x |
| Brilliant Violet 785™ anti-human CD4 Antibody | BV785 | CD4 | OKT4 | BioLegend | 317441 | B426990 | 400x |
| FITC anti-human HLA-DR | FITC | HLA-DR | L243 | BioLegend | 980402 | B326372 | 300x |
| PerCP/Cyanine5.5 anti-human CD39 Antibody | PerCP-Cy5.5 | CD39 | A1 | BioLegend | 328217 | B339378 | 200x |
| PE anti-human FOXP3 Recombinant Antibody | PE | Foxp3 | QA18A03 | BioLegend | 364703 | B444720 | 400x |
| PE/Dazzle™ 594 anti-human CD3 Antibody | Pe-Dazzle594 | CD3 | OKT3 | BioLegend | 317345 | B443448 | 400x |
| PE/Cyanine7 anti-human CD45RA Antibody | PE-Cy7 | CD45RA | HI100 | BioLegend | 304125 | B426645 | 400x |
| APC anti-human GM-CSF Antibody | APC | GM-CSF | BVD2-21C11 | BioLegend | 502309 | B327459 | 100x |
| Alexa Fluor® 700 anti-human Perforin Antibody | AF700 | PRF1 | B-D48 | BioLegend | 353323 | B332512 | 400x |
| Zombie NIR Fixable Viability Kit | NIR | Via |  | BioLegend | 423106 | B419021 | 1000x |
| **Panel 3 - Activated cells** |  |  |  |  |  |  |  |
| BD Horizon™ BUV395 Mouse Anti-Human CD8 | BUV395 | CD8a | RPA-T8 | BD | 563795 | 1097745 | 400x |
| BD Horizon™ BUV737 Mouse Anti-Human CD69 | BUV737 | CD69 | FN50 | BD | 612817 | 2095735 | 400x |
| BD Horizon™ BUV805 Mouse Anti-Human CD14 | BUV805 | CD14 | M5E2 | BD | 612903 | 4250797 | 400x |
| CellTrace™ Violet Cell Proliferation Kit | BV421 | CTV |  | ThermoFisher | C34571 |  | 1000x |
| Brilliant Violet 605™ anti-human CD25 Antibody | BV605 | CD25 | BC96 | BioLegend | 302631 | B355185 | 400x |
| BD Horizon™ BV711 Mouse Anti-Human IFN-γ | BV711 | IFNγ | B27 | BD | 564039 | 1197770 | 300x |
| Brilliant Violet 785™ anti-human CD4 Antibody | BV785 | CD4 | OKT4 | BioLegend | 317441 | B426990 | 400x |
| FITC anti-human/mouse Granzyme B Antibody | FITC | GZMB | GB11 | BioLegend | 515403 | B351810 | 300x |
| BD Pharmingen™ PerCP-Cy™5.5 Mouse Anti-Human IL-17A | PerCP-Cy5.5 | IL17A | N49-653 | BD | 560799 | 1131141 | 100x |
| PE anti-human T-bet Recombinant Antibody | PE | Tbet | QA18A24 | BioLegend | 365903 | B412345 | 300x |
| PE/Dazzle™ 594 anti-human CD3 Antibody | Pe-Dazzle594 | CD3 | OKT3 | BioLegend | 317345 | B443448 | 400x |
| PE/Cyanine7 anti-human Granzyme A Antibody | PE-Cy7 | GZMA | CB9 | BioLegend | 507221 | B360334 | 100x |
| APC anti-human GM-CSF Antibody | APC | GMCSF | BVD2-21C11 | BioLegend | 502309 | B327459 | 100x |
| Alexa Fluor® 700 anti-human Perforin Antibody | AF700 | PRF1 | B-D48 | BioLegend | 353323 | B332512 | 400x |
| Zombie NIR Fixable Viability Kit | NIR | Via |  | BioLegend | 423106 | B419021 | 1000x |
| **Sort** |  |  |  |  |  |  |  |
| Brilliant Violet 421™ anti-human CD4 Antibody | BV421 | CD4 | OKT4 | BioLegend | 317433 | B431672 | 200x |
| Zombie Aqua™ Fixable Viability Kit | BV510 | Via |  | BioLegend | 423101 |  | 1000x |
| FITC anti-human CD186 (CXCR6) Antibody | FITC | CXCR6 | K041E5 | BioLegend | 356019 | B358258 | 100x |
| PE anti-human CD27 Antibody | PE | CD27 | M-T271 | BioLegend | 356405 | B350974 | 200x |
| PE/Cyanine7 anti-human CD8 Antibody | PE-Cy7 | CD8a | SK1 | BioLegend | 344711 | B431460 | 200x |
| APC/Cyanine7 anti-human CD3 Antibody | APC-Cy7 | CD3 | HIT3a | BioLegend | 300317 | B289411 | 200x |
| Human TruStain FcX™ (Fc Receptor Blocking Solution) | TruStain FcX |  |  |  | 422302 | B365487 | 500x |

**Supplementary Table S5.** Reagents used for flow cytometry of murine cells.

| Name | Fluorochrome | Marker | Clone | Manufacturer | Cat |
| --- | --- | --- | --- | --- | --- |
| APC/Cyanine7 anti-mouse TCR $\beta$ chain Antibody | APC-Cy7 | TCRβ | H57-597 | BioLegend | 109220 |
| APC anti-mouse CD4 Antibody | APC | CD4 | GK1.5 | BioLegend | 100412 |
| FITC anti-mouse CD8a Antibody | FITC | CD8α | 53-6.7 | BioLegend | 100706 |
| PE/Cyanine7 anti-mouse CD8b Antibody | PE-Cy7 | CD8β | YTS156.7.7 | BioLegend | 126616 |
| PE anti-mouse CD62L Antibody | PE | CD62L | MEL-14 | BioLegend | 104408 |
| Pacific Blue™ anti-human/mouse Granzyme B Recombinant Antibody | PB | GZMB | QA16A02 | BioLegend | 372218 |
| Alexa Fluor® 594 anti-rat CD90/mouse CD90.1 (Thy1.1) Antibody | AF594 | CD90.1 | OX-7 | BioLegend | 202545 |
| Brilliant Violet 785™ anti-mouse CD90.2 (Thy1.2) Antibody | BV785 | CD90.2 | 30-H12 | BioLegend | 105331 |
| Fixable Viability Dye eFluor™ 506 | EF506 | Viability |  | Invitrogen | 65-0866-14 |

## Notes

### Author Declarations

Ethical protocols were approved by the Fred Hutchinson Cancer Center Institutional Review Board (Protocol #9730), the Boston Children's Hospital Institutional Review Board (Protocol #IRB-P00030890), and the Seattle Children's Institutional Review Board (Protocol #1638).

## References

1 Hracs, L. et al. Global evolution of inflammatory bowel disease across epidemiologic stages. Nature, doi:10.1038/s41586-025-08940-0 (2025).

2 Papamichael, K. et al. Role for Therapeutic Drug Monitoring During Induction Therapy with TNF Antagonists in IBD: Evolution in the Definition and Management of Primary Nonresponse. Inflammatory Bowel Diseases 21, 182–197, doi:10.1097/MIB.0000000000000202 (2015).

3 Gibble, T. H. et al. Identification of inadequate responders to advanced therapy among commercially-insured adult patients with Crohn’s disease and ulcerative colitis in the United States. BMC gastroenterology 23, 63, doi:10.1186/s12876-023-02675-w (2023).

4 Smillie, C. S. et al. Intra- and Inter-cellular Rewiring of the Human Colon during Ulcerative Colitis. Cell 178, 714–730.e722, doi:10.1016/j.cell.2019.06.029 (2019).

5 Martin, J. C. et al. Single-Cell Analysis of Crohn’s Disease Lesions Identifies a Pathogenic Cellular Module Associated with Resistance to Anti-TNF Therapy. Cell 178, 1493–1508.e1420, doi:10.1016/j.cell.2019.08.008 (2019).

6 Thomas, T. et al. A longitudinal single-cell atlas of anti-tumour necrosis factor treatment in inflammatory bowel disease. Nature Immunology 25, 2152–2165, doi:10.1038/s41590-024-01994-8 (2024).

7 Jostins, L. et al. Host–microbe interactions have shaped the genetic architecture of inflammatory bowel disease. Nature 491, 119–124, doi:10.1038/nature11582 (2012).

8 Liu, J. Z. et al. Association analyses identify 38 susceptibility loci for inflammatory bowel disease and highlight shared genetic risk across populations. Nature Genetics 47, 979–986, doi:10.1038/ng.3359 (2015).

9 Goyette, P. et al. High-density mapping of the MHC identifies a shared role for HLA-DRB1*01:03 in inflammatory bowel diseases and heterozygous advantage in ulcerative colitis. Nature Genetics 47, 172–179, doi:10.1038/ng.3176 (2015).

10 Biton, M. et al. T Helper Cell Cytokines Modulate Intestinal Stem Cell Renewal and Differentiation. Cell 175, 1307–1320.e1322, doi:10.1016/j.cell.2018.10.008 (2018).

11 Brabec, T. et al. Segmented filamentous bacteria-induced epithelial MHCII regulates cognate CD4+ IELs and epithelial turnover. The Journal of experimental medicine 221, doi:10.1084/jem.20230194 (2024).

12 Heuberger, C. E. et al. MHC class II antigen presentation by intestinal epithelial cells fine-tunes bacteria-reactive CD4 T-cell responses. Mucosal Immunology 17, 416–430, 10.1016/j.mucimm.2023.05.001 (2024).

13 Ciccarelli, F., De Martinis, M. & Ginaldi, L. An update on autoinflammatory diseases. Current medicinal chemistry 21, 261–269, doi:10.2174/09298673113206660303 (2014).

14 Parsa, R. et al. Newly recruited intraepithelial Ly6A^+^CCR9^+^CD4^+^ T&#xa0;cells protect against enteric viral infection. Immunity 55, 1234–1249.e1236, doi:10.1016/j.immuni.2022.05.001 (2022).

15 Mortha, A. et al. Neutralizing Anti-Granulocyte Macrophage-Colony Stimulating Factor Autoantibodies Recognize Post-Translational Glycosylations on Granulocyte Macrophage-Colony Stimulating Factor Years Before Diagnosis and Predict Complicated Crohn&#x2019;s Disease. Gastroenterology 163, 659–670, doi:10.1053/j.gastro.2022.05.029 (2022).

16 Uzzan, M. et al. Ulcerative colitis is characterized by a plasmablast-skewed humoral response associated with disease activity. Nat Med 28, 766–779, doi:10.1038/s41591-022-01680-y (2022).

17 Yokoi, T. et al. Identification of a unique subset of tissue-resident memory CD4^+^ T cells in Crohn&#x2019;s disease. Proceedings of the National Academy of Sciences 120, e2204269120, doi:doi:10.1073/pnas.2204269120 (2023).

18 Arase, M. et al. Multi-omics uncovers transcriptional programs of gut-resident memory CD4+ T cells in Crohn’s disease. Journal of Experimental Medicine 222, doi:10.1084/jem.20242106 (2025).

19 Quandt, J. et al. Wnt–β-catenin activation epigenetically reprograms Treg cells in inflammatory bowel disease and dysplastic progression. Nature Immunology 22, 471–484, doi:10.1038/s41590-021-00889-2 (2021).

20 Martini, G. R. et al. Selection of cross-reactive T cells by commensal and food-derived yeasts drives cytotoxic TH1 cell responses in Crohn’s disease. Nature Medicine 29, 2602–2614, doi:10.1038/s41591-023-02556-5 (2023).

21 Elmentaite, R. et al. Cells of the human intestinal tract mapped across space and time. Nature 597, 250–255, doi:10.1038/s41586-021-03852-1 (2021).

22 Zheng, H. B. et al. (eLife Sciences Publications, Ltd, 2023).

23 Nettey, L. et al. Divergent T Cell Phenotypes Define Pediatric Crohn’s Disease and Ulcerative Colitis. medRxiv, 2025.2010.2016.25336112, doi:10.1101/2025.10.16.25336112 (2025).

24 Sinopoulou, V. et al. Immunomodulators and Advanced Therapies for Induction of Remission in Crohn’s Disease: A Systematic Review and Network Meta-Analysis. Inflammatory Bowel Diseases 32, 53–66, doi:10.1093/ibd/izaf191 (2025).

25 Geremia, A., Biancheri, P., Allan, P., Corazza, G. R. & Di Sabatino, A. Innate and adaptive immunity in inflammatory bowel disease. Autoimmunity reviews 13, 3–10, doi:10.1016/j.autrev.2013.06.004 (2014).

26 Andreatta, M. & Carmona, S. J. STACAS: Sub-Type Anchor Correction for Alignment in Seurat to integrate single-cell RNA-seq data. Bioinformatics 37, 882–884, doi:10.1093/bioinformatics/btaa755 (2021).

27 Love, M. I., Huber, W. & Anders, S. Moderated estimation of fold change and dispersion for RNA-seq data with DESeq2. Genome Biol 15, 550, doi:10.1186/s13059-014-0550-8 (2014).

28 Pariente, B. et al. Activation of the Receptor NKG2D Leads to Production of Th17 Cytokines in CD4^+^ T Cells of Patients With Crohn’s Disease. Gastroenterology 141, 217–226.e212, doi:10.1053/j.gastro.2011.03.061 (2011).

29 Riaz, T., Sollid, L. M., Olsen, I. & de Souza, G. A. Quantitative Proteomics of Gut-Derived Th1 and Th1/Th17 Clones Reveal the Presence of CD28+ NKG2D- Th1 Cytotoxic CD4+ T cells*. Molecular & Cellular Proteomics 15, 1007–1016, 10.1074/mcp.M115.050138 (2016).

30 Kazer, S. W. et al. Primary nasal influenza infection rewires tissue-scale memory response dynamics. Immunity 57, 1955–1974.e1958, 10.1016/j.immuni.2024.06.005 (2024).

31 Street, K. et al. Slingshot: cell lineage and pseudotime inference for single-cell transcriptomics. BMC genomics 19, 477, doi:10.1186/s12864-018-4772-0 (2018).

32 Roux de Bézieux, H., Van den Berge, K., Street, K. & Dudoit, S. Trajectory inference across multiple conditions with condiments. Nat Commun 15, 833, doi:10.1038/s41467-024-44823-0 (2024).

33 Nandy, A. et al. Epstein-Barr Virus Exposure Precedes Crohn’s Disease Development. Gastroenterology 169, 150–153, doi:10.1053/j.gastro.2025.01.247 (2025).

34 Read, K. A. et al. Aiolos represses CD4+ T cell cytotoxic programming via reciprocal regulation of TFH transcription factors and IL-2 sensitivity. Nature Communications 14, 1652, doi:10.1038/s41467-023-37420-0 (2023).

35 Jones, D. M. et al. Cytotoxic Programming of CD4+ T Cells Is Regulated by Opposing Actions of the Related Transcription Factors Eos and Aiolos. Journal of immunology (Baltimore, Md. : 1950) 212, 1129–1141, doi:10.4049/jimmunol.2300748 (2024).

36 Eshima, K. et al. Ectopic expression of a T-box transcription factor, eomesodermin, renders CD4(+) Th cells cytotoxic by activating both perforin- and FasL-pathways. Immunology letters 144, 7–15, doi:10.1016/j.imlet.2012.02.013 (2012).

37 Knudson Cory, J., et al. Mechanisms of Antiviral Cytotoxic CD4 T Cell Differentiation. Journal of Virology 95, 10.1128/jvi.00566-00521, doi:10.1128/jvi.00566-21 (2021).

38 Patil, V. S. et al. Precursors of human CD4^+^ cytotoxic T lymphocytes identified by single-cell transcriptome analysis. Science Immunology 3, eaan8664, doi:doi:10.1126/sciimmunol.aan8664 (2018).

39 Müller-Dott, S. et al. Expanding the coverage of regulons from high-confidence prior knowledge for accurate estimation of transcription factor activities. Nucleic Acids Research 51, 10934–10949, doi:10.1093/nar/gkad841 (2023).

40 Sang-aram, C., Browaeys, R., Seurinck, R. & Saeys, Y. Unraveling cell–cell communication with NicheNet by inferring active ligands from transcriptomics data. Nature Protocols 20, 1439–1467, doi:10.1038/s41596-024-01121-9 (2025).

41 Neurath, M. F. Cytokines in inflammatory bowel disease. Nature Reviews Immunology 14, 329–342, doi:10.1038/nri3661 (2014).

42 Gerhardt, L., Hong, M. M. Y., Yousefi, Y., Figueredo, R. & Maleki Vareki, S. IL-12 and IL-27 Promote CD39 Expression on CD8+ T Cells and Differentially Regulate the CD39+CD8+ T Cell Phenotype. Journal of immunology (Baltimore, Md. : 1950) 210, 1598–1606, doi:10.4049/jimmunol.2200897 (2023).

43 Mascanfroni, I. D. et al. IL-27 acts on DCs to suppress the T cell response and autoimmunity by inducing expression of the immunoregulatory molecule CD39. Nat Immunol 14, 1054–1063, doi:10.1038/ni.2695 (2013).

44 Bréart, B. et al. IL-27 elicits a cytotoxic CD8+ T cell program to enforce tumour control. Nature 639, 746–753, doi:10.1038/s41586-024-08510-w (2025).

45 Peters, A. et al. IL-27 Induces Th17 Differentiation in the Absence of STAT1 Signaling. Journal of immunology (Baltimore, Md. : 1950) 195, 4144–4153, doi:10.4049/jimmunol.1302246 (2015).

46 Lucas, S., Ghilardi, N., Li, J. & de Sauvage, F. J. IL-27 regulates IL-12 responsiveness of na&#xef;ve CD4^+^ T cells through Stat1-dependent and -independent mechanisms. Proceedings of the National Academy of Sciences 100, 15047–15052, doi:doi:10.1073/pnas.2536517100 (2003).

47 Martin, E. et al. Role of IL-27 in Epstein–Barr virus infection revealed by IL-27RA deficiency. Nature 628, 620–629, doi:10.1038/s41586-024-07213-6 (2024).

48 Kar, R., Sinha, S., Khatun, Z., Sharma, A. & Patil, V. S. A distinct subset of stem-cell memory is poised for the cytotoxicity program in CD4^+^ T cells in humans. Science Advances 12, eady6423, doi:doi:10.1126/sciadv.ady6423 (2026).

49 Yang, Y. et al. Focused specificity of intestinal TH17 cells towards commensal bacterial antigens. Nature 510, 152–156, doi:10.1038/nature13279 (2014).

50 Ivanov, II et al. Induction of intestinal Th17 cells by segmented filamentous bacteria. Cell 139, 485–498, doi:10.1016/j.cell.2009.09.033 (2009).

51 Paprckova, D. et al. Self-reactivity of CD8 T-cell clones determines their differentiation status rather than their responsiveness in infections. Frontiers in Immunology Volume 13 - 2022, doi:10.3389/fimmu.2022.1009198 (2022).

52 Hao, Y. et al. Integrated analysis of multimodal single-cell data. Cell 184, 3573–3587.e3529, doi:10.1016/j.cell.2021.04.048 (2021).

53 Preglej, T. & Ellmeier, W. CD4(+) Cytotoxic T cells - Phenotype, Function and Transcriptional Networks Controlling Their Differentiation Pathways. Immunology letters 247, 27–42, doi:10.1016/j.imlet.2022.05.001 (2022).

54 Oh, D. Y. & Fong, L. Cytotoxic CD4(+) T cells in cancer: Expanding the immune effector toolbox. Immunity 54, 2701–2711, doi:10.1016/j.immuni.2021.11.015 (2021).

55 Hasegawa, T. et al. Cytotoxic CD4^+^ T&#xa0;cells eliminate senescent cells by targeting cytomegalovirus antigen. Cell 186, 1417–1431.e1420, doi:10.1016/j.cell.2023.02.033 (2023).

56 Balasubramanian, P. et al. Single-cell RNA profiling of blood CD4+ T cells identifies distinct helper and dysfunctional regulatory clusters in children with SLE. Nature Immunology 26, 2100–2111, doi:10.1038/s41590-025-02297-2 (2025).

57 Cachot, A. et al. Tumor-specific cytolytic CD4 T cells mediate immunity against human cancer. Science Advances 7, eabe3348, doi:10.1126/sciadv.abe3348 (2021).

58 Hashimoto, K. et al. Single-cell transcriptomics reveals expansion of cytotoxic CD4 T cells in supercentenarians. Proceedings of the National Academy of Sciences of the United States of America 116, 24242–24251, doi:10.1073/pnas.1907883116 (2019).

59 Ma, J. et al. Single-cell analysis pinpoints distinct populations of cytotoxic CD4(+) T cells and an IL-10(+)CD109(+) T(H)2 cell population in nasal polyps. Sci Immunol 6, doi:10.1126/sciimmunol.abg6356 (2021).

60 Soghoian, D. Z. et al. HIV-Specific Cytolytic CD4 T Cell Responses During Acute HIV Infection Predict Disease Outcome. Science Translational Medicine 4, 123ra125–123ra125, doi:10.1126/scitranslmed.3003165 (2012).

61 Jamwal, D. R. et al. Intestinal Epithelial Expression of MHCII Determines Severity of Chemical, T-Cell&#x2013;Induced, and Infectious Colitis in Mice. Gastroenterology 159, 1342–1356.e1346, doi:10.1053/j.gastro.2020.06.049 (2020).

62 Katsanos, K. H., Papamichael, K., Feuerstein, J. D., Christodoulou, D. K. & Cheifetz, A. S. Biological therapies in inflammatory bowel disease: Beyond anti-TNF therapies. Clinical Immunology 206, 9–14, 10.1016/j.clim.2018.03.004 (2019).

63 Berg, D. R., Colombel, J.-F. & Ungaro, R. The Role of Early Biologic Therapy in Inflammatory Bowel Disease. Inflammatory Bowel Diseases 25, 1896–1905, doi:10.1093/ibd/izz059 (2019).

64 Wang, H. et al. Type I interferon drives T cell cytotoxicity by upregulation of interferon regulatory factor 7 in autoimmune kidney diseases in mice. Nature Communications 16, 4686, doi:10.1038/s41467-025-59819-7 (2025).

65 Venzin, V. & Iannacone, M. Reframing IL-27: a central regulator of CD8(+) T cell immunity. Trends in immunology, doi:10.1016/j.it.2025.10.012 (2025).

66 Bhat, P., Leggatt, G., Waterhouse, N. & Frazer, I. H. Interferon-γ derived from cytotoxic lymphocytes directly enhances their motility and cytotoxicity. Cell death & disease 8, e2836, doi:10.1038/cddis.2017.67 (2017).

67 McGregor, C. et al. Spatial fibroblast niches define Crohn’s fistulae. Nature, doi:10.1038/s41586-025-09744-y (2025).

68 Kong, L. et al. Single-cell and spatial transcriptomics of stricturing Crohn’s disease highlights a fibrosis-associated network. Nature Genetics 57, 1742–1753, doi:10.1038/s41588-025-02225-y (2025).

69 Aran, D. et al. Reference-based analysis of lung single-cell sequencing reveals a transitional profibrotic macrophage. Nat Immunol 20, 163–172, doi:10.1038/s41590-018-0276-y (2019).

70 Mabbott, N. A., Baillie, J. K., Brown, H., Freeman, T. C. & Hume, D. A. An expression atlas of human primary cells: inference of gene function from coexpression networks. BMC genomics 14, 632, doi:10.1186/1471-2164-14-632 (2013).

71 Niederlova, V. et al. Imbalance of stem-like and effector T cell states in children with early type 1 diabetes across conventional and regulatory subsets. Nature Communications 16, 11301, doi:10.1038/s41467-025-66459-4 (2025).

72. Korotkevich, G., et al. Fast gene set enrichment analysis. 060012, doi: 10.1101/060012% J bioRxiv (2021).

73 Neavin, D. et al. Demuxafy: improvement in droplet assignment by integrating multiple single-cell demultiplexing and doublet detection methods. Genome Biology 25, 94, doi:10.1186/s13059-024-03224-8 (2024).

74 Mombaerts, P. et al. RAG-1-deficient mice have no mature B and T lymphocytes. Cell 68, 869–877, doi:10.1016/0092-8674(92)90030-g (1992).

